# Evaluation of plasma p-tau217 for detecting amyloid pathology in a diverse and heterogeneous community-based cohort

**DOI:** 10.1101/2025.01.20.25320851

**Authors:** Marc D. Rudolph, Courtney L. Sutphen, Thomas C. Register, Samuel N. Lockhart, Melissa M. Rundle, Timothy M. Hughes, James R. Bateman, Kiran K. Solingapuram Sai, Christopher T. Whitlow, Suzanne Craft, Michelle M. Mielke

## Abstract

**INTRODUCTION:** Studies suggest excellent performance of plasma p-tau217 for detecting amyloid pathology, though studies in more diverse populations are needed to validate previously determined cutpoints.

**METHODS:** Plasma p-tau217 utility for detecting amyloid pathology (Aβ) via amyloid PET (*n*=598) and/or cerebrospinal fluid (CSF; *n*=154) was assessed in a heterogeneous, community-based cohort in the Wake Forest Alzheimer’s Disease Research Center (WFADRC). Participants (*n*=598) were 21% Black; 313 cognitive unimpaired (CU), 214 mild cognitive impairment (MCI), and 64 dementia (DEM); 49% prediabetic, 44% hypertensive; 29% overweight/obese; and 64% with mild-to-moderate kidney disease. Gaussian-mixture models, logistic regression, and receiver operating curve analyses were performed.

**RESULTS:** Plasma p-tau217 was associated with elevated Aβ deposition and accurately classified Aβ-positive participants (PET: AUC: 94%-97%, cutpoint≥.338 pg/mL; CSF: AUC = .84, cutpoint ≥.307 pg/mL).

**DISCUSSION:** Plasma p-tau217 is an accurate indicator of amyloid pathology in a heterogeneous cohort, and superior to other plasma biomarkers assessed. Longitudinal analyses assessing impact of comorbidities on p-tau217 utility for disease progression are underway.

**HIGHLIGHTS:**

- The WFADRC is a diverse and heterogeneous cohort.
- P-tau217 levels were lower, on average, in cognitively unimpaired participants, females, and Black participants.
- Plasma p-tau217 classified amyloid PET positive individuals with high precision and performed better than p-tau181.
- Cutpoints and reference ranges of plasma p-tau217 were lower compared to recently published thresholds.
- Combining cutpoint approaches, a 4-tier system captured cohort heterogeneity.

## 1 BACKGROUND

Plasma biomarkers are a minimally invasive and cost-effective means of detecting Alzheimer’s disease (AD) pathology [1–3]. Plasma phosphorylated tau 217 (p-tau217) has been shown to be a sensitive peripheral marker of brain amyloid plaque deposition [4–15]. Recent studies suggest excellent performance of plasma ALZpath p-tau217 for detecting elevated amyloid PET [10,16,17]. However, studies incorporating more *heterogeneous* populations are needed to further validate the previously examined cutpoints and develop a range of biomarker thresholds.

As previously described [1,7,18], comorbidities such as BMI and chronic kidney disease (CKD) can influence plasma biomarker levels due to physiological reasons (i.e. renal function or blood volume) and which may result in false positive diagnoses of AD pathology [19]. Additional metabolic factors such as insulin resistance and hypertension have been linked to altered amyloid levels as detected by PET [20–23]. Importantly, these comorbidities have a higher prevalence among underrepresented minorities, but few studies have examined the accuracy of ALZpath p-tau217 in diverse and heterogeneous community cohorts where multiple comorbidities may be present [19].

Using data from the Wake Forest Alzheimer’s Disease Research Center (WFADRC), we comprehensively examined the utility of plasma p-tau217 for detecting amyloid pathology (Aβ) detected in-vivo using both positron emission tomography (PET) and cerebrospinal fluid (CSF) among a diverse and heterogeneous community-based cohort. We also considered the impact of chronic disease and other health factors at baseline. Participants in the WFADRC (*n* = 598) were adjudicated as either cognitively unimpaired (52.51%; *n* = 314), having mild cognitive impairment (35.62%; *n* = 213), dementia (10.7%; *n* = 64), or other impairment not specified (<1 %; *n* = 7). From the overall cohort, 21% were Black/African American, 12% with CKD (64% with early-stage kidney disease), 12% diabetic (49% prediabetic), 29% overweight or obese; and 43% were hypertensive.

## 2 METHODS & MATERIALS

### 2.1 Participants

All participants were enrolled in the WFADRC, a cohort that includes cognitively unimpaired participants as well as those in the early stages of cognitive impairment and dementia. As previously described, adults between the ages of 55 and 85 were recruited from the surrounding community and by referral through our memory clinic into the WFADRC (2016-2022) via the WFADRC Clinical Core supported by efforts of the Outreach, Engagement & Recruitment Core [1,24]. Participants underwent a standard evaluation, including the National Alzheimer’s Coordinating Center (NACC) protocol for clinical research data collection, clinical exams, neurocognitive testing, neuroimaging, and genotyping for the apolipoprotein E (*APOE*) ε4 allele. Exclusion criteria included a history of large vessel stroke (participants with lacunae or small vessel ischemic disease were eligible); other significant neurologic diseases; uncontrolled *chronic* medical or psychiatric conditions (such as advanced liver or severe kidney disease [eGFR < 30]; poorly controlled congestive heart failure, chronic obstructive pulmonary disease or sleep apnea; active cancer treatment; uncontrolled clinical depression, or psychiatric illness; current use of insulin; and history of substance abuse or heavy alcohol consumption within the previous ten years). The Wake Forest Institutional Review Board approved all activities as described; written informed consent was obtained for all participants and/or their legally authorized representatives.

### 2.2 Adjudication

An expert panel of investigators including neuropsychologists, neurologists, and geriatricians, provided adjudication of cognitive status. Following determination of clinical cognitive diagnosis for cognitively unimpaired (CU), mild cognitive impairment (MCI) [25], dementia (DEM) [26], and other impairment not specified (OTHER), adjudication of cause/type of cognitive impairment or dementia was assessed using neuroimaging and fluid biomarkers.

#### 2.4.4. Assessment of demographics and health conditions

Age, race/ethnicity (e.g., Black/White; Non-Hispanic), sex (e.g., male/female), and years of education were self-reported. *APOE* genotype was obtained by Taqman using single nucleotide polymorphisms (rs429358 and rs7412) to determine haplotypes of ε2, ε3, and ε4. *APOE* was dichotomized to represent the presence or absence of one or more ε4 alleles (e.g., carrier vs. non-carrier; *APOE*-*ε*4). Total body-mass index (BMI) was calculated as weight (lb)/[height (in)]^2^ x 703. Kidney function was evaluated in blood from individuals free from severe CKD using the estimated glomerular filtration rate (eGFR; severe [Stage G4-G5; National Kidney Foundation] CKD defined as eGFR < 30; also see participant characteristics) via Labcorp assay presented in units of mL/min/1.73m^2. Fasting blood glucose levels were measured via an oral glucose tolerance test (OGTT) from serial blood draws; impaired glucose tolerance was defined by glucose ≥ 140 mg/dL at 120 minutes of OGTT or hemoglobin A1c ≥ 5.7% as previously described [27]. Diabetic status (normal; prediabetic; diabetic) was determined using a combination of OGTT tests (either baseline or 120) and hemoglobin A1c levels. Specifically, participants were coded as diabetic if A1C ≥ 6.5, OGTT (baseline) ≥ 126, or OGTT (120 minutes) ≥ 200, or as prediabetic if meeting the following criteria 5.7 < A1c < 6.5 or 100 < OGTT (baseline) < 126 or 140 < OGTT (120 minutes) < 200 (all remaining participants were coded as normal). Hypertension status (HTN) was defined according to 2017 ACC/AHA guidelines as seated blood pressure ≥ 130 mmHg, diastolic blood pressure ≥ 80 mmHg, and/or current use of antihypertensive medications as described elsewhere [1]. Cardiometabolic index (CMI) was calculated using the product of two ratios (waist/height x triglycerides/HDL) [28]. Participant medical histories including an active or recent history of stroke (CBSTROKE), traumatic brain injury (TBI), or myocardial infarction (MYOINF) were captured using the Uniform Data Set Version 3 (UDSv3) [29].

### 2.4 Biomarkers

#### 2.4.1 MRI Imaging

Participants were scanned on a research-dedicated 3-Tesla Siemens Skyra magnetic resonance imaging (MRI; 32-channel head coil) scanner. Detailed image acquisition parameters have been previously published [24,27].

#### 2.4.2 Aβ-PET Imaging

As previously described [1,30] fibrillar Aβ brain deposition on PET was assessed with [^11^C]-Pittsburgh compound B (PiB) [31]. Following a computed tomography (CT) scan for attenuation correction, participants were injected with an intravenous bolus of ∼10mCi (approximately 370 MBq) PiB and scanned from 40–70 minutes (6×5-min frames) post-injection on a 64-slice GE Discovery MI DR PET/CT scanner. Each participant’s CT image was coregistered to their structural MRI, and PET frames were coregistered to MRI space using the affine matrix from the CT-MRI coregistration. SUVr images were co-registered and resliced to the T1 structural MRI closest in time within ∼6 months on average (Months: M=.98; SD = 5.9). Aβ deposition was quantified using a voxelwise standardized uptake volume ratio (SUVr), calculated as the PiB SUVr (40-70 min, cerebellar grey reference) signal averaged from a cortical meta-ROI sensitive to the early pathogenesis of AD relative to the uptake in the cerebellum, using FreeSurfer-segmented (v7.2; https://surfer.nmr.mgh.harvard.edu) regions [30,32]. Centiloid (CL) values were also calculated to facilitate harmonization across ligands and centers [33]. CL analysis was conducted in PMOD v4.1 (PMOD LLC Technologies, Switzerland). The 3D T1-weighted MRI and aligned average PET scan were input into the PMOD PNEURO Step-wise Maximum Probability Atlas workflow using the Centiloid atlas template validated using the Global Alzheimer’s Association Interactive Network (GAAIN) data set. Briefly, MRIs were normalized to the MNI-space template and segmented. Coregistered PETs were normalized to MNI space using MRI parameters. A PMOD SUVr was calculated using the standard MNI-space Centiloid ROI with the whole cerebellum as a reference region. Centiloid scores were then calculated using Klunk et al. equation 1.3b. CL =100(pibSUVrIND-1.009)/1.067. Visual reads were conducted by (MMR, SNL, and JRB) following published criteria [34,35].

Global PiB (Aβ-PET; [FreeSurfer] SUVr and CL) served as a biomarker of amyloid burden used in all analyses. Amyloid positivity (Aβ− & Aβ+) was assessed using a combination of (1) visual reads, (2) several previously defined thresholds set *a priori* for both SUVr and Centiloid scales (SUVr ≥ 1.21; CL ≥ 12 and CL ≥ 24) [36–38], and (3) and a sample-specific data-driven threshold (CL ≥ 32). We report results for all amyloid PET thresholds assessed. However, we primarily focus on CL ≥ 24 to define amyloid PET positivity to be consistent with recently published work assessing the ALZpath p-tau217 assay [10].

#### 2.4.3 Plasma and Fluid Biomarkers

Plasma and CSF biomarkers were collected from subjects after a minimum 8 hour (water only) fast and processed according to National Centralized Repository for Alzheimer’s Disease (NCRAD) protocols.

##### 2.4.3.1 Plasma biomarkers

Blood was processed within 30 minutes of collection as described previously [1]. For plasma, EDTA-treated tubes were inverted 10 times and placed on wet ice before centrifugation at 2000g at 4°C for 15 minutes. Processed plasma was then aliquoted into polypropylene tubes and stored at −80°C until analysis. Plasma p-tau217 samples, collected from 2017-2023, were processed in duplicate using ALZpath Simoa p-tau 217 v2 assay kits on a Quanterix HD-X at Neurocode (Bellingham WA). Duplicates with CVs>20% or missing one value were repeated. Kit QC controls were run with each plate. The coefficient of variation (CV; Mean = 4.47; SD = 3.64) for p-tau217 was well within previously established reference ranges (maximum CV of 20) and in-line with other cohorts as recently published [10]. All samples were processed in duplicate per the manufacturer’s instructions. For plasma p-tau181, analyses were conducted at NCRAD as previously described [1] Plasma pTau181 was assessed using the Quanterix SIMOA pTau181 version 2 Advantage Kits. Blood samples and PET data were acquired at different timepoints; thus, we evaluated if the time interval between blood collection and PET acquisition at baseline impacted relationships between plasma p-tau217 and Aβ deposition or Aβ positivity.

##### 2.4.3.2 CSF biomarkers

Following local anesthesia, a 22-gauge Sprotte needle was used to perform lumbar puncture (LP) for the collection of CSF by the gravity method from the L3-4 (∼80%) or L4-5 (∼20%) interspace while participants rested in the seated or lateral decubitus position. CSF was transferred into pre-chilled low protein binding Sarstedt polypropylene tubes (Cat. 72.703.600) and stored frozen at −80°C until time of analysis. CSF levels of Aβ42, Aβ40, total tau, and phospho-tau (pTau181) were measured from 1st thawed samples using a Lumipulse G1200 Chemiluminescence Enzyme Immunoassay (CLEIA) System and reagents and calibrators from the manufacturer (Fujirebio Diagnostics Inc., Malvern, PA). Performance of kit controls are as follows, for AB42, AB40, and ptau181, intraassay CVs were < 3.1%, for total tau the CVs were <5.1%. The ratio of Aβ42/Aβ40 was used to define CSF amyloid positivity in our cohort [39,40]. Amyloid positivity was determined using the Aβ42-to Aβ40 ratio (Aβ42/40) from CSF using an in-house threshold (CSF Aβ42/40 ≤ .058) [41,42].

### 2.5 Statistical Analysis

Raw unadjusted and Winsorized Pearson and Spearman correlation coefficients are provided in the supplemental material for all primary continuous measures of interest. General linear models assessed associations between p-tau217 and amyloid PET positivity before and after covariate adjustment (Model 1 [unadjusted]: Model 2 [adjusting for age, sex, race, education, APOE status, BMI, and eGFR]: Model 3 [Model 2 covariates plus cognitive status, diabetes group, hypertension, TBI, stroke, and myocardial infarction]). All variables were z-transformed prior to analysis using multivariable logistic regression. Sample-derived (e.g., data-driven; Aβ-PET agnostic) positivity cutoffs were derived using gaussian-mixture modeling (GMM) [36,43,44]. GMMs were constructed using the *normalmixEM* function from the *mixtools* R package [45]. Receiver-operating curve (ROC) analyses were used to derive cutpoints for p-tau217 that maximized concordance with Aβ-PET positivity estimates (SUVr & CL). Cutpoints were derived using the *cutpointr* function from the *Cutpointr* R package [46] specifying 10,000 bootstrap iterations and maximizing the sum of sensitivity and specificity (e.g., Youden index). Two-point detection thresholds (e.g., two cutoff model) maximizing sensitivity and specificity at the lower and upper bound of the ROC 95% confidence interval, hereafter referred to as max-sensitivity and max-specificity, were generated using *pROC* R package [47] as previously described [10]. For all models, we report the area-under-the-receiver-operating curve (AUC) and their respective bootstrapped confidence intervals, sensitivity and specificity, positive and negative predictive values, average accuracy, and prevalence of a given outcome (e.g., amyloid positivity). Confidence intervals for the optimal cutpoints and AUC values were extracted using the *boot_ci* function. Non-parametric Wilcoxon signed rank tests, were performed to provide effect size (ES) estimates comparing biomarkers levels after stratifying the sample by clinical diagnosis, sex, race, *APOE*-ε4 carriership, diabetic status, and CKD. Corresponding effect sizes were reported to aid with interpretation when there are a limited number of observations (e.g. race). Effect sizes were interpreted using established guidelines (Wilcoxon *r*: negligible < .10, small (S) = .10-.30, moderate (M) = .30-.50, large (L) ≥ .50; Cohen’s *d*: negligible < .20, small < .50, medium < .80; large ≥ .80) [48]. Multiple comparisons corrections were performed using the Benjamini-Hochberg false-discovery rate (*p*-FDR) [49]. All analyses were conducted in R (RStudio Team, 2020).

## 3 RESULTS

### 3.1 Participants

Participant characteristics are provided in Table 1. The mean (standard deviation, SD) age of participants at baseline was 69.89 ± 7.99; 21% self-identified as Black/African American; 65% as female; and 32% were *APOE* ε4 carriers. Approximately 71% of participants had *mildly decreased* eGFR (60-90) and 15% met criteria for monitoring of early-stage kidney disease (eGFR < 60) [50,51]. 60% of participants had or were at risk for diabetes at their baseline visit (11.5% diabetic; 48.5% prediabetic), 44% were hypertensive, and 29% were classified as either obese or overweight. Approximately 34% were amyloid PET positive at baseline (CL ≥ 24; see Table S1 for a characteristic breakdown of the cohort stratified by amyloid PET positivity).

**Table 1.**
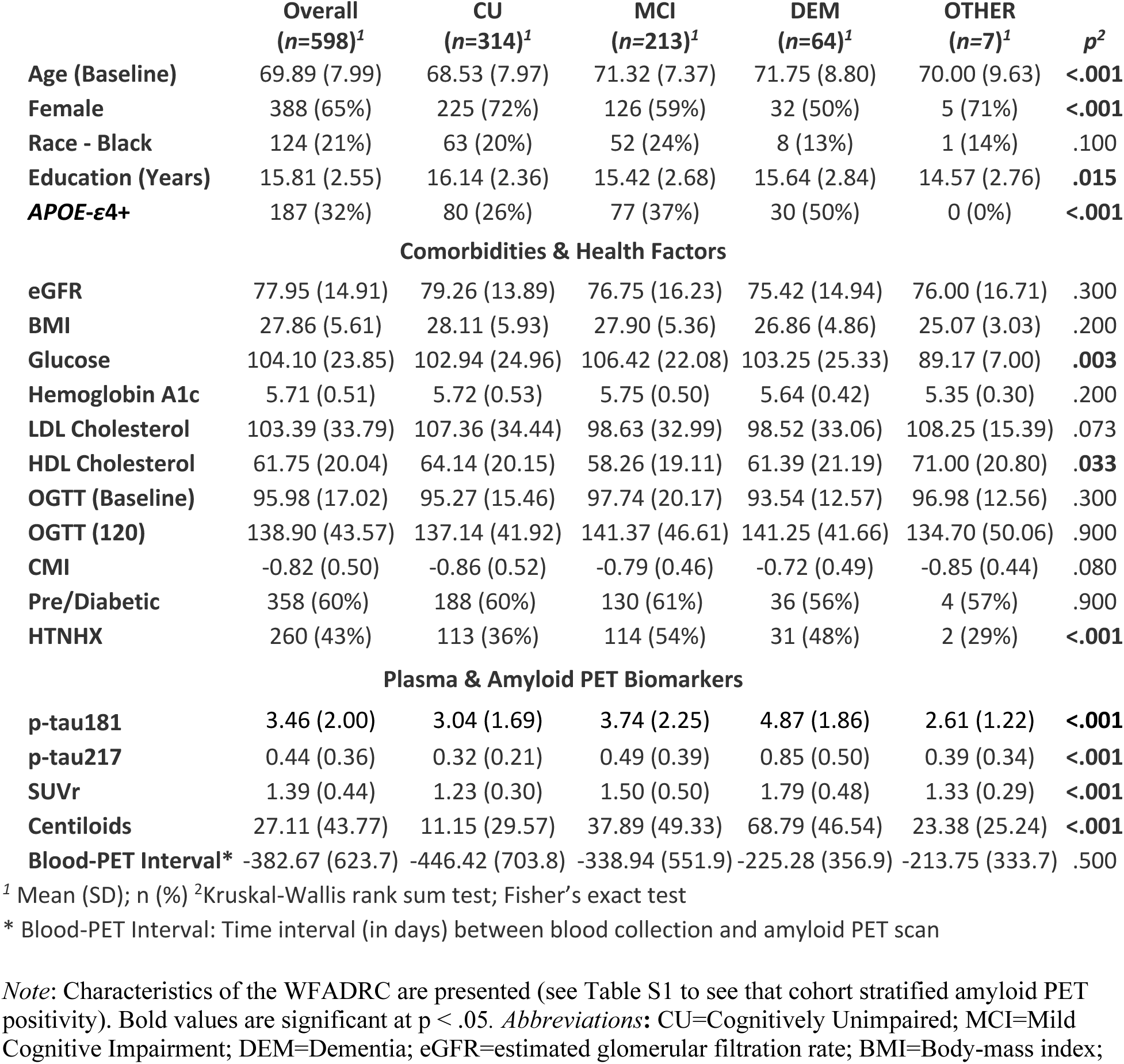
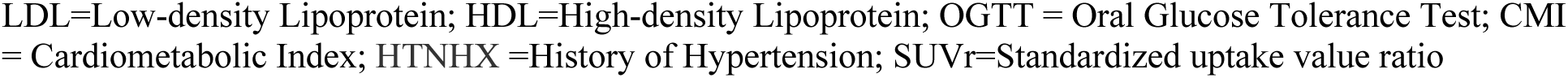
Participant Characteristics.

|  | **Overall**<br>**(n=598)<sup>1</sup>** | **CU**<br>**(n=314)<sup>1</sup>** | **MCI**<br>**(n=213)<sup>1</sup>** | **DEM**<br>**(n=64)<sup>1</sup>** | **OTHER**<br>**(n=7)<sup>1</sup>** | **p<sup>2</sup>** |
| --- | --- | --- | --- | --- | --- | --- |
| **Age (Baseline)** | 69.89 (7.99) | 68.53 (7.97) | 71.32 (7.37) | 71.75 (8.80) | 70.00 (9.63) | **<.001** |
| **Female** | 388 (65%) | 225 (72%) | 126 (59%) | 32 (50%) | 5 (71%) | **<.001** |
| **Race - Black** | 124 (21%) | 63 (20%) | 52 (24%) | 8 (13%) | 1 (14%) | .100 |
| **Education (Years)** | 15.81 (2.55) | 16.14 (2.36) | 15.42 (2.68) | 15.64 (2.84) | 14.57 (2.76) | **.015** |
| **APOE-ε4+** | 187 (32%) | 80 (26%) | 77 (37%) | 30 (50%) | 0 (0%) | **<.001** |
| **Comorbidities & Health Factors** |  |  |  |  |  |  |
| **eGFR** | 77.95 (14.91) | 79.26 (13.89) | 76.75 (16.23) | 75.42 (14.94) | 76.00 (16.71) | .300 |
| **BMI** | 27.86 (5.61) | 28.11 (5.93) | 27.90 (5.36) | 26.86 (4.86) | 25.07 (3.03) | .200 |
| **Glucose** | 104.10 (23.85) | 102.94 (24.96) | 106.42 (22.08) | 103.25 (25.33) | 89.17 (7.00) | **.003** |
| **Hemoglobin A1c** | 5.71 (0.51) | 5.72 (0.53) | 5.75 (0.50) | 5.64 (0.42) | 5.35 (0.30) | .200 |
| **LDL Cholesterol** | 103.39 (33.79) | 107.36 (34.44) | 98.63 (32.99) | 98.52 (33.06) | 108.25 (15.39) | .073 |
| **HDL Cholesterol** | 61.75 (20.04) | 64.14 (20.15) | 58.26 (19.11) | 61.39 (21.19) | 71.00 (20.80) | **.033** |
| **OGTT (Baseline)** | 95.98 (17.02) | 95.27 (15.46) | 97.74 (20.17) | 93.54 (12.57) | 96.98 (12.56) | .300 |
| **OGTT (120)** | 138.90 (43.57) | 137.14 (41.92) | 141.37 (46.61) | 141.25 (41.66) | 134.70 (50.06) | .900 |
| **CMI** | -0.82 (0.50) | -0.86 (0.52) | -0.79 (0.46) | -0.72 (0.49) | -0.85 (0.44) | .080 |
| **Pre/Diabetic** | 358 (60%) | 188 (60%) | 130 (61%) | 36 (56%) | 4 (57%) | .900 |
| **HTNHX** | 260 (43%) | 113 (36%) | 114 (54%) | 31 (48%) | 2 (29%) | **<.001** |
| **Plasma & Amyloid PET Biomarkers** |  |  |  |  |  |  |
| **p-tau181** | 3.46 (2.00) | 3.04 (1.69) | 3.74 (2.25) | 4.87 (1.86) | 2.61 (1.22) | **<.001** |
| **p-tau217** | 0.44 (0.36) | 0.32 (0.21) | 0.49 (0.39) | 0.85 (0.50) | 0.39 (0.34) | **<.001** |
| **SUVr** | 1.39 (0.44) | 1.23 (0.30) | 1.50 (0.50) | 1.79 (0.48) | 1.33 (0.29) | **<.001** |
| **Centiloids** | 27.11 (43.77) | 11.15 (29.57) | 37.89 (49.33) | 68.79 (46.54) | 23.38 (25.24) | **<.001** |
| **Blood-PET Interval\*** | -382.67 (623.7) | -446.42 (703.8) | -338.94 (551.9) | -225.28 (356.9) | -213.75 (333.7) | .500 |

<sup>1</sup> Mean (SD); n (%) <sup>2</sup>Kruskal-Wallis rank sum test; Fisher's exact test
\* Blood-PET Interval: Time interval (in days) between blood collection and amyloid PET scan
*Note:* Characteristics of the WFADRC are presented (see Table S1 to see that cohort stratified amyloid PET positivity). Bold values are significant at $p < .05$ . *Abbreviations:* CU=Cognitively Unimpaired; MCI=Mild Cognitive Impairment; DEM=Dementia; eGFR=estimated glomerular filtration rate; BMI=Body-mass index;
LDL=Low-density Lipoprotein; HDL=High-density Lipoprotein; OGTT = Oral Glucose Tolerance Test; CMI = Cardiometabolic Index; HTNHX =History of Hypertension; SUVr=Standardized uptake value ratio

### 3.2 Primary Analyses

#### 3.2.1 General Linear Models

Plasma p-tau217 levels were elevated in: (1) males compared to females (0.470 vs. 0.419; *p* = .006; ES = .109); (2) White versus Black participants (0.467 vs. 0.331, p < .001; ES=0.196); and (3) in *APOE*-ε4 carriers vs. non-carriers (0.541 vs. 0.379, p < .001; ES=0.236). Baseline plasma p-tau217 was positively correlated with age (*rho* =.36, *p* < .001) and negatively correlated with eGFR (*rho* = −.23, *p* < .001) and with BMI (*rho* = −.16, *p* < .001). Baseline plasma p-tau217 and p-tau181 were positively correlated (*rho*=.75; *p* < .001). Baseline plasma p-tau181 and p-tau217 were positively correlated with Aβ-PET deposition (Figure 1; p-tau181 [Figure 1, panel a]: overall *rho*=.49, *p* < .001; p-tau217 [Figure 1, panel b]: overall *rho* =.70, p< .001).

**Figure 1.**
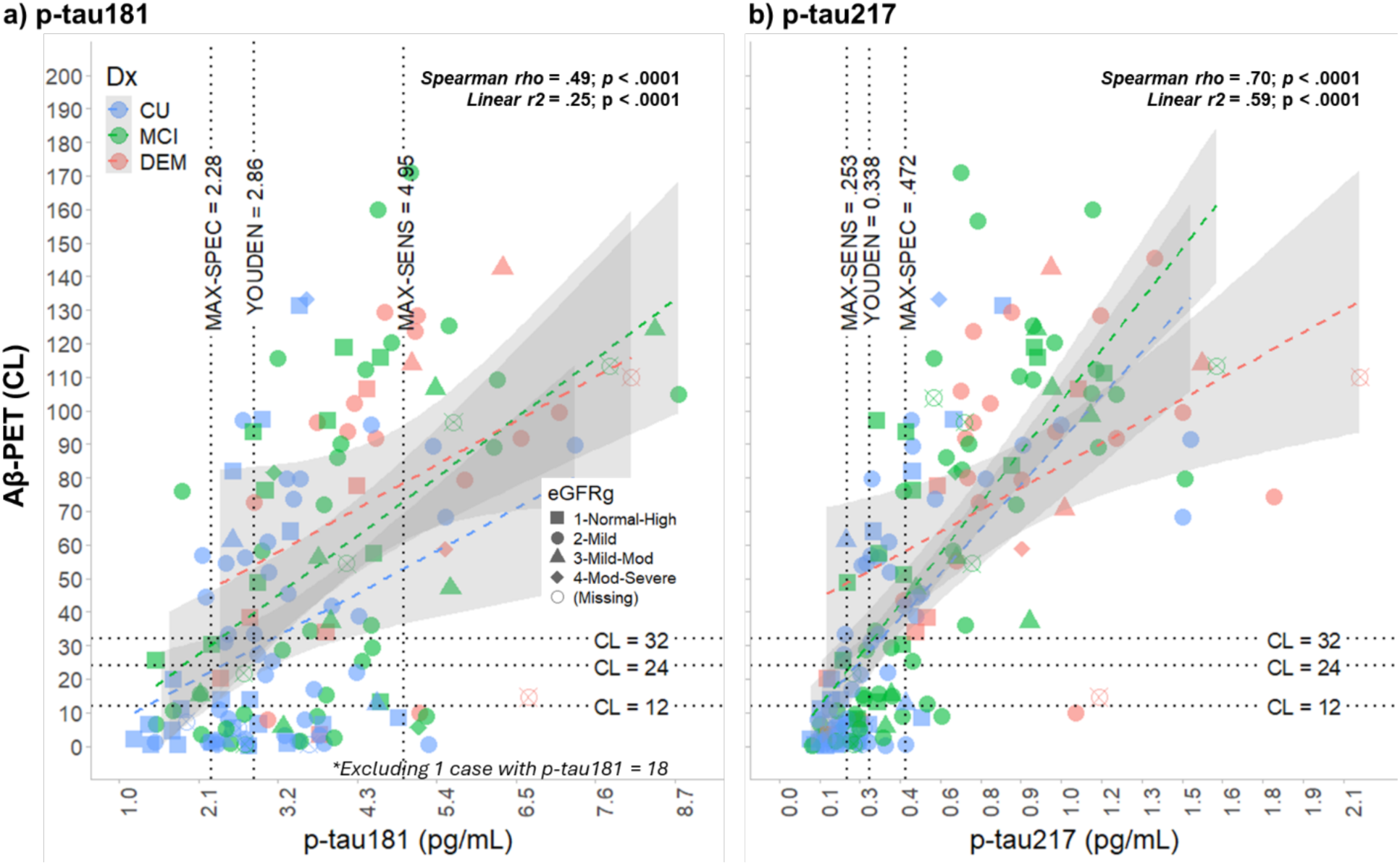
Associations between amyloid PET deposition (Centiloids) and plasma p-tau181 and p-tau217. *Note:* Associations between amyloid PET deposition (Centiloids) and plasma p-tau181 (panel a) and p-tau217(panel b). Scatterplots highlight a stronger correlation between p-tau217 and amyloid PET deposition across cognitive status groups compared to p-tau181 and amyloid PET. Dotted lines represent optimal (binary) and two-point detection cutpoints for each p-tau variable observed when classifying amyloid PET positivity. Data points are color-coded by diagnosis and of different shapes based on eGFR status (Stage 1 [Normal High] eGFR >= 90; Stage 2 [Mild] > 60 eGFR < 90; Stage 3 [Mild-Moderate] > 45 eGFR < 60; Stage 4 [Moderate Severe] eGFR < 45); *Abbreviations*: CU = Cognitively Unimpaired; MCI = Mild Cognitive Impairment; DEM = Dementia; CL = Centiloids; SENS = Sensitivity; SPEC = Specificity; eGFRg = Estimated Glomerular Filtration Rate.

Baseline plasma p-tau181 and p-tau217 were positively correlated with Aβ-PET positivity (e.g., higher in Aβ-positive participants) here defined as CL ≥ 24 before and after covariate adjustment (all *p* < .05; Table 2). For p-tau181, several covariates significantly contributed to model performance (Model 2: Age and *APOE*-ε4 carriership; Model 3: Age, sex, race, *APOE*-ε4 carriership, and diagnostic status; all *p*-FDR < .05). For p-tau217, sex was significant prior to FDR correction (*p* = .031) in the fully adjusted model only (Model 3). All other covariates *p* ≥ .05 before and after FDR correction.

**Table 2.**
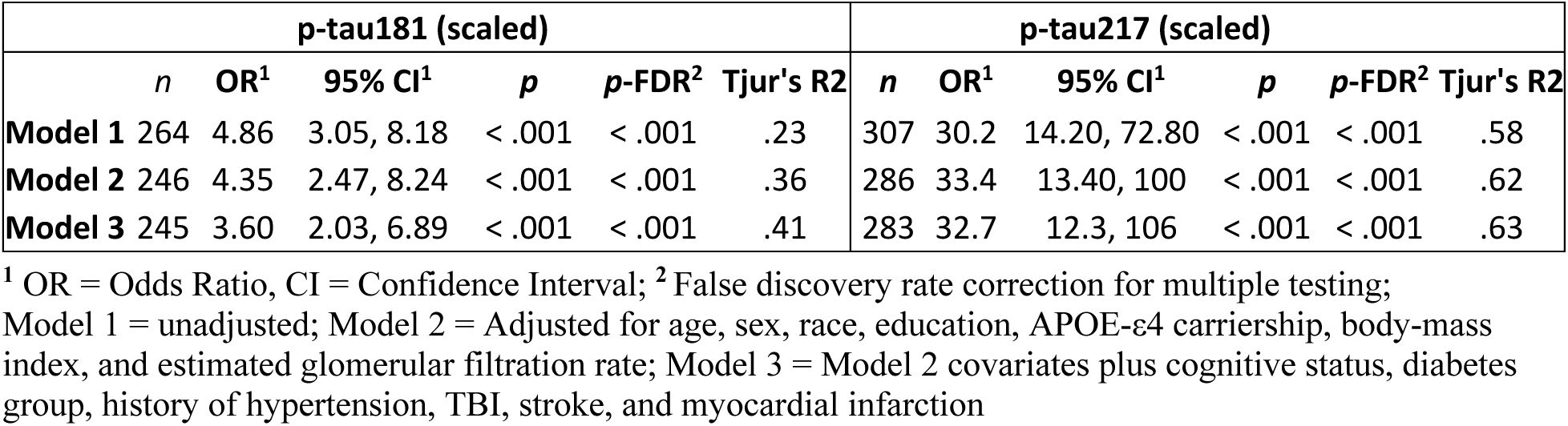
Logistic regression results assessing associations between amyloid PET positivity (CL ≥ 24) and plasma p-tau181 and p-tau217.

| p-tau181 (scaled) |  |  |  |  |  |  | p-tau217 (scaled) |  |  |  |  |  |  |
| --- | --- | --- | --- | --- | --- | --- | --- | --- | --- | --- | --- | --- | --- |
|  | *n* | OR<sup>1</sup> | 95% CI<sup>1</sup> | *p* | *p*-FDR<sup>2</sup> | Tjur's R<sup>2</sup> |  | *n* | OR<sup>1</sup> | 95% CI<sup>1</sup> | *p* | *p*-FDR<sup>2</sup> | Tjur's R<sup>2</sup> |
| **Model 1** | 264 | 4.86 | 3.05, 8.18 | < .001 | < .001 | .23 |  | 307 | 30.2 | 14.20, 72.80 | < .001 | < .001 | .58 |
| **Model 2** | 246 | 4.35 | 2.47, 8.24 | < .001 | < .001 | .36 |  | 286 | 33.4 | 13.40, 100 | < .001 | < .001 | .62 |
| **Model 3** | 245 | 3.60 | 2.03, 6.89 | < .001 | < .001 | .41 |  | 283 | 32.7 | 12.3, 106 | < .001 | < .001 | .63 |

<sup>1</sup> OR = Odds Ratio, CI = Confidence Interval; <sup>2</sup> False discovery rate correction for multiple testing; Model 1 = unadjusted; Model 2 = Adjusted for age, sex, race, education, APOE- ε4 carriership, body-mass index, and estimated glomerular filtration rate; Model 3 = Model 2 covariates plus cognitive status, diabetes group, history of hypertension, TBI, stroke, and myocardial infarction

#### 3.2.2 Classification & Cutpoint Analysis

Table 3 and Figure 2 depicts cutpoints for both plasma p-tau181 and p-tau217, derived using receiver operating curve analyses and gaussian-mixture models, and include optimal and two-point ROC detection cutpoints. Positive and negative predictive values ranged from .75-.84 for p-tau181 and .89-.97 for p-tau217. Additional details and a full table of classification results across amyloid PET thresholds can be found in Table S2. Plasma p-tau181 classified Aβ-PET positivity (AUC: 77-82%) across thresholds (Figure 2, panel a;). Baseline plasma p-tau217 accurately classified Aβ-PET positivity (Table 3, Figure 2, panel b; AUC: 91-96%) across thresholds.

**Figure 2.**
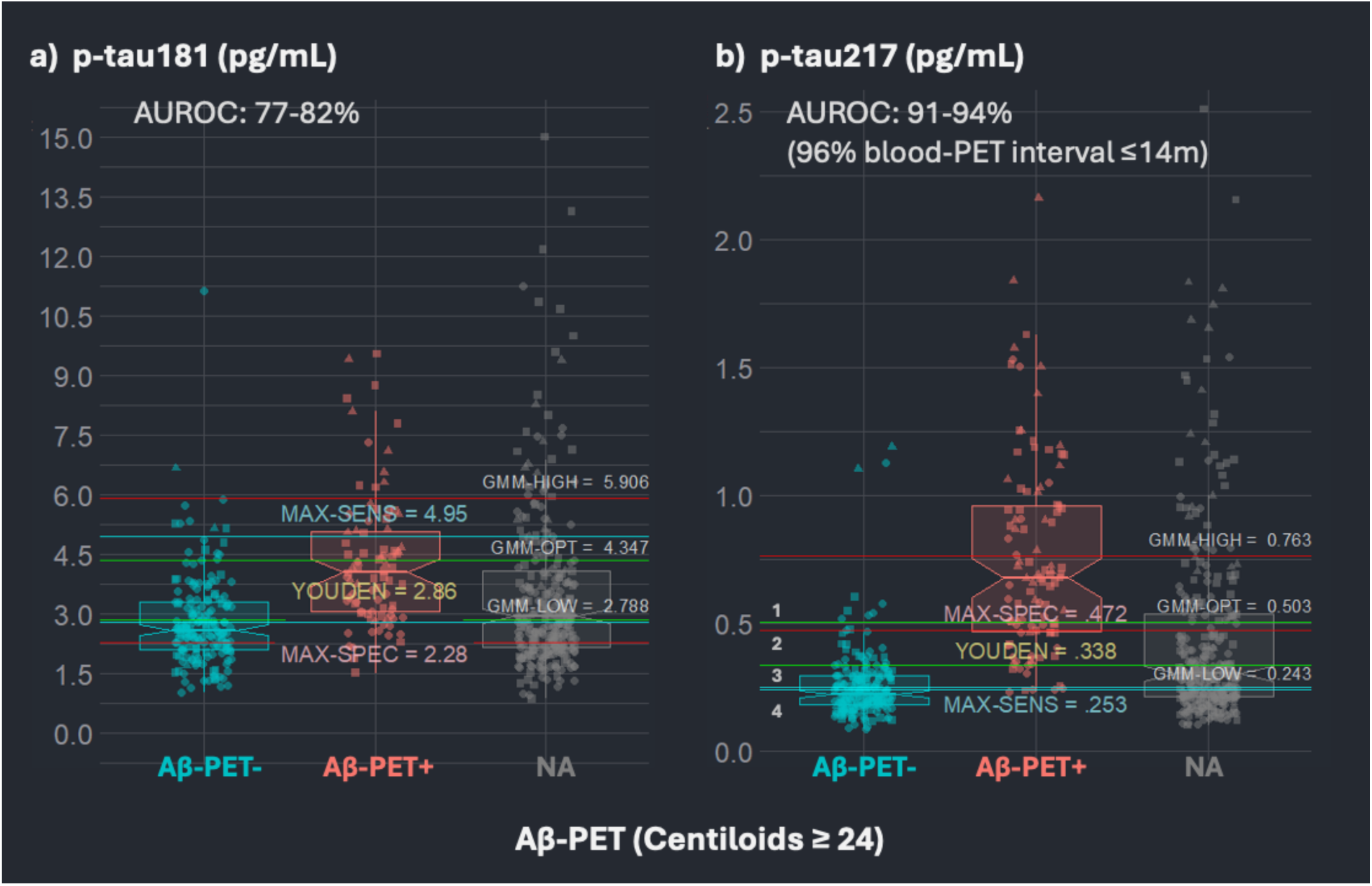
Performance of p-tau181 and p-tau217 for classifying amyloid PET positivity defined as ≥ 24 Centiloids. *Note:* Figure 2 depicts cutpoints derived using receiver operating curve analyses and gaussian-mixture models for both plasma (a) p-tau181 and (b) p-tau217. Optimal (e.g., binary; Youden index = .338 pg/mL) and two-point ROC detection thresholds (MAX-SENS=.253 pg/mL; MAX-SPEC=.472 pg/mL) were combined resulting in a 4-tier system: (1) Negative [<0.253; N∼39%]; (2) Intermediate-Low [0.253-.338; N∼20%]; (3) Intermediate-High [0.338-.472; 13%]; and (4) Positive [>0.472; 28%]. *Abbreviations*: Aβ = Amyloid Beta; CL = Centiloids; SENS = Sensitivity; SPEC = Specificity; GMM = Gaussian-Mixture Models; NA = Missing (e.g., Plasma p-tau217 datapoints without a corresponding amyloid PET scan).

**Table 3.**
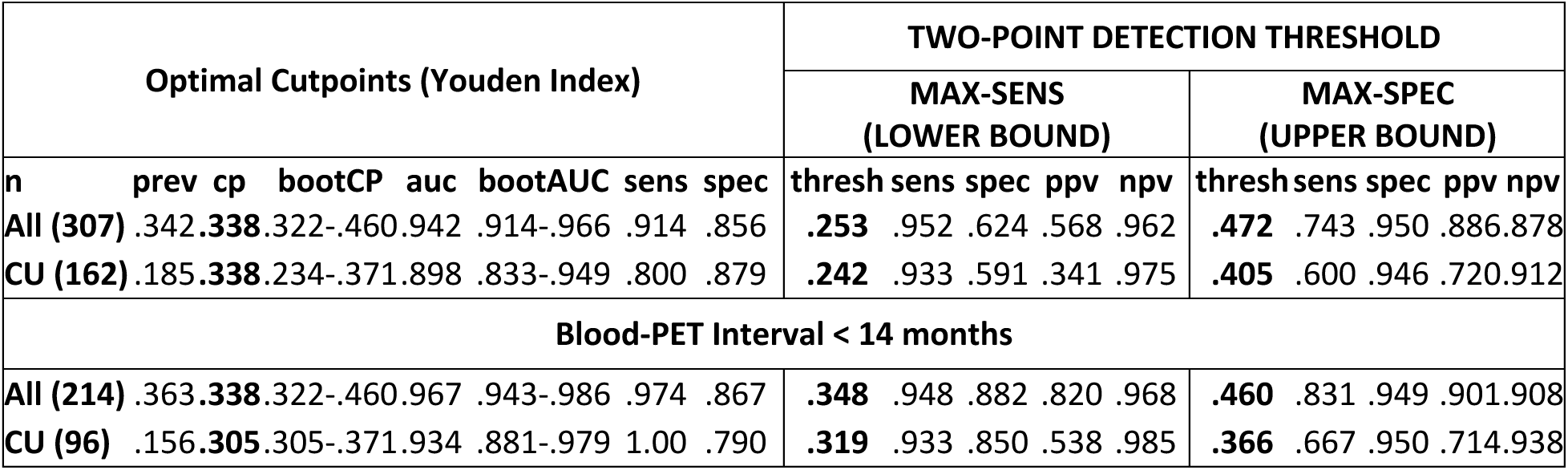
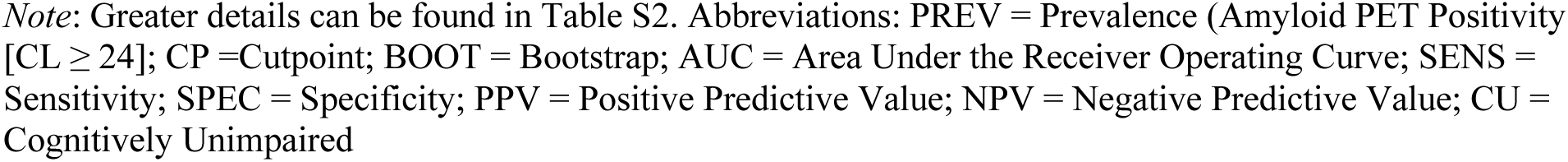
Optimal cutpoints and two-point detection thresholds for p-tau217 when classifying amyloid PET positivity (≥ 24 Centiloids)

For p-tau217, combining optimal (Youden index = .338 pg/mL) and two-point detection thresholds (max-sensitivity = .253; max-specificity = .472) stratified the cohort into four zones: (1) Negative [<.253; *n*=241 (40%)]; (2) Intermediate-Low [.253-.338; *n*=109 (18%)]; (3) Intermediate-High [.338-.472; *n*=77 (13%)]; and (4) Positive [>.472; *n*=171 (29%)]). Descriptive statistics by these four zones are provided in Table S3. GMM-based cutpoints based on the distribution of observed plasma p-tau217 values were higher than those reported above for classifying amyloid PET positivity in the full sample and when restricting the sample to CU participants (see Figure S1).

##### 3.2.2.1 Blood-PET Interval

The average time between the date of plasma collection and amyloid PET scan acquisition was 305 days (SD = 605 days; Figure S2). Approximately 75% of the cohort had their first PET scan within 1-2 years of their initial blood draw (< 1 year: 209 (67%), 1-2 years: 21 (6.7%), 2-3 years; 34 (11%); 3-4 years: 23 (7.3%); > 4 years: 26 (8.3.4%). The time between the date of blood collection and PET scan acquisition was greater than 1 year for 15 of the 38 (∼40%) discordant cases identified (see Supplemental Materials Section 2.2 for additional details).

The time interval between blood collection and initial amyloid pet scan did not correlate with plasma or neuroimaging biomarker levels (p-tau217: rho= −.08, *p* = .128; SUVr: rho= −.06, *p* = .992; CL: rho= −.01, *p* = .797) and did not differ by either cognitive status or amyloid PET positivity when stratifying the cohort by sex, race, *APOE*, diabetes or hypertension, or kidney functioning (all *p* > .05). Excluding participants with intervals greater than or less than 14 months increased the precision of p-tau217 to classify Aβ-PET+ across thresholds evaluated (AUC: Visual Read = 97%; SUVr ≥ 1.21 = 93%; CL ≥ 12 = 96%; CL ≥ 24 = 97%; CL ≥ 32 = 97%). The optimal binary cutpoint remained consistent at .338 pg/mL.

##### 3.2.2.2 Discordance

There were 38 participants classified as discordant between the plasma and PET measure (p-tau217+/Aβ-PET-: n=29; p-tau217-/Aβ-PET+: n=9). High discordance values were defined using the upper (p-tau217 ≥ .472 pg/mL/Aβ-PET-: *n*=10) and lower (p-tau217 ≤ .253 pg/mL/Aβ-PET+: *n*=5) two-point detection points. Discordance across amyloid PET thresholds is provided in Table S4. Descriptive characteristics for discordant cases, before and after excluding cases based on the time between blood collection and initial PET scan, are provided in Table S5 and Table S6. Briefly, we observed an interesting pattern such that participants with higher baseline levels of p-tau217 who were Aβ-PET negative (p-tau217+/AB-PET-) were more likely to have a history of hypertension, whereas the few participants with low p-tau217 levels considered Aβ-PET positive (p-tau217-/AB-PET+) had poorer cardiometabolic health, were Black, and female.

#### 3.2.3 CSF-derived amyloid positivity

We further evaluated p-tau217 cutpoints using CSF positivity in a smaller subset of participants (*n*=154). The performance of plasma p-tau217 was slightly lower (AUC=.844) and more variable when classifying amyloid positivity defined using CSF compared to Aβ-PET (Table 4). Likewise, binary (e.g., Youden) cutpoints were similar, albeit lower, to the cutpoints derived using Aβ-PET in our cohort given the smaller sample size and prevalence of Aβ+ individuals.

**Table 4.** Performance of ALZpath p-tau217 to classify amyloid positivity defined by CSF Aβ42/40.

| CSF | prev | cp | bootCP | AUC | bootAUC | sens | spec | acc |
| --- | --- | --- | --- | --- | --- | --- | --- | --- |
| All ($n = 154$) | .41 | 0.307 | 0.248-0.535 | .844 | .780-.903 | .810 | .747 | .770 |
| CU ($n=96$) | .29 | 0.307 | 0.234-0.361 | .727 | .594-.847 | .783 | .783 | .750 |

*Note:* Optimal cutpoints obtained using an in-house cutpoint (CSF Aβ + < .058). *Abbreviations:* CSF = Cerebrospinal Fluid; CP = Cutpoint; Boot = Bootstrapped; AUC = Area Under the Receiver Operating Curve; SENS = Sensitivity; SPEC = Specificity; ACC = Accuracy; PREV = Prevalence; CU = Cognitively Unimpaired

#### 3.2.3 Cognitive Status

As in Table 1, p-tau217 significantly differed on average between CU, MCI, and dementia groups (*p* < .05). In terms of classification accuracy, p-tau217 best distinguished CU vs DEM participants (AUC=83%; CU vs MCI: AUC=63%; MCI vs DEM: AUC=73%). Results were comparable after excluding large blood-PET intervals (CU vs MCI: AUC=69%; CU vs DEM: AUC=83%; MCI vs DEM: AUC=70%).

#### 3.2.4 Sensitivity Analyses

Several additional sensitivity analyses were conducted. Table S7 examined p-tau217 cutpoints before and after excluding large intervals between blood collection and scan acquisition (see Table 1 for cognitive status). Figures S3-S7 provide optimal cutpoints of p-tau217 when stratified by clinical diagnosis, sex, race, and APOE. Overall, we observed similar performance when classifying amyloid PET positivity across the various strata. Correspondingly, marginal group differences were observed for plasma p-tau217 and amyloid PET deposition when stratifying the cohort based on sex, race, and kidney function (see Tables S11-S12 and Figures S3-S7).

## 4 DISCUSSION

Our primary finding is that plasma p-tau217, measured with the ALZpath antibody, is strongly associated with elevated amyloid PET deposition and accurately classified amyloid positive individuals in a diverse and heterogenous cohort. This finding is consistent with a recent study from a more homogenous cohort (Ashton et al., 2024). However, the cutpoint that we identified was lower. In addition, and similar to other studies, we also found that p-tau217 was superior to p-tau181 for classifying amyloid PET positivity across diagnostic groups, and was largely independent of demographic factors, comorbidities, and other health-related factors (Benedet et al., 2022; Janelidze et al., 2023; Mielke et al., 2022; Milà-Alomà et al., 2022). Ashton and colleagues (2024) previously assessed the utility of the ALZpath p-tau217 to classify amyloid-PET positivity across three independent cohorts including Translational Biomarkers in Aging and Dementia (TRIAD), Wisconsin Registry for Alzheimer’s Prevention (WRAP), and the Sant Pau Initiative on Neurodegeneration (SPIN). Although the mean ALZpath p-tau217 levels obtained in the current cohort were comparable to the WRAP cohort, they were lower than those obtained in the TRIAD and SPIN cohorts. In addition, the cutpoint derived in the current cohort (Youden index = .338 pg/mL) was lower that the cutpoints previously identified by Ashton and colleagues (positive: >0.63 pg/mL; intermediate: 0.4 – 0.63; pg/mL; negative: <0.4 pg/mL) [10].

Several factors must be considered when comparing absolute cutoffs between cohorts. First, the prevalence of Aβ-PET+ in the WFADRC was approximately 32%-42% across the various amyloid PET thresholds examined. This is a higher prevalence than that reported by Ashton et al. in the WRAP cohort (21% were Aβ-PET+ (CL≥ 24)) Second, we observed lower plasma p-tau217 cutpoints regardless of how Aβ-PET positivity was defined and when stratifying the cohort by cognitive status, sex, race, *APOE*-*ε*4 carriership, and either hypertensive or diabetic status. Restricting our cohort to cognitively unimpaired participants with amyloid PET with blood-PET interval < 14 months, resulted in the lowest optimal cutpoint (p-tau217 ≥ .305 pg/mL).

Second, the recruitment methods and cohort characteristics can impact the cutpoint. As background, the WFADRC focuses on the transition from normal aging to MCI and then to AD and ADRD, with emphasis on understanding the contribution of metabolic and vascular factors to these transitions. Recruitment efforts target cognitively unimpaired or MCI patients with special emphasis on recruitment of persons from underrepresented groups. In general, the cohort matches the racial, ethnic, comorbidity, educational and sociodemographic characteristics of the surrounding community. As part of the adjudication process, cases with clinical/biomarker discordance or severity were often referred for additional assessments. These recruitment decisions may skew the cohort distribution of PET biomarker data points, and thus affect positivity thresholds. As noted in the introduction, 48% of the WFADRC cohort is pre-diabetic and 46% is hypertensive. Black participants make up ∼21% of the cohort assessed here and tended to have poorer cardiometabolic health on average as here indexed by the cardiometabolic index (waist/height x triglycerides/HDL). Thus, it remains crucial to investigate multiple biomarker positivity thresholds to robustly assess overall positivity in diverse settings and cohorts.

Third, the observed cutpoints may differ between cohorts due to natural variation in biomarker levels. That is, though beyond the scope of this paper, additional factors pertaining to the collection, storage, and processing of blood or plasma samples may prove to be critical if the ultimate goal is to determine a set of universal cutpoints [52,53] that generalize across cohorts and real-world samples. In the present study, plasma p-tau217 sample were assayed with the same platform at the same time from all participants. Ongoing harmonization efforts and standardized protocols are likely to partially help mitigate any *cross-cohort* discrepancies in detected plasma levels and cutpoints [52,54–56]. Our results suggest thresholds for p-tau217, based on PET and CSF biomarkers, should consider what impact covariates such as age, sex, race, cognitive status, and overall health including kidney functioning have on cohort-specific biomarker levels.

The prevalence of amyloid positivity directly impacts biomarker classification performance and the number of specific cases that may be considered discordant at any given timepoint. Therefore, in the present study we considered multiple established and cohort-specific amyloid PET thresholds. However, differences in tracers used to quantify amyloid PET deposition also warrant consideration. A small study reported near-identical performance when comparing 11C-PiB (utilized in the current study) and 18F-AZD4694 (TRIAD, WRAP, and SPIN cohorts) to detect cortical amyloid [57]. Thus, while the specific tracer may not largely influence a single cutpoint, it remains unknown whether specific tracers perform differently in the presence of multiple comorbidities and health-related risk factors Therefore, it remains critical for studies to (1) recruit and retain participants from underrepresented groups; (2) ensure robust reporting of cohort demographics and health histories, especially kidney function, and (3) provide detailed methodological reporting (e.g., biomarker collection, storage, processing, handling, and timing) [5,52,54]. Likewise, in our cohort, p-tau217 better classified amyloid positivity defined by amyloid PET as compared to CSF Aβ42/40, which may be attributed to the restricted CSF-sample, however, recent work suggests PET may better capture earlier brain Aβ deposition [58].

We also observed that plasma p-tau217 performed relatively worse when attempting to distinguish participants based on cognitive status. When the goal is to screen a participant for potential treatment with an anti-amyloid monoclonal antibody therapy, a single cutpoint is desirable, however, health-related risk factors need to be considered, in particular when cases are just above or below a specified cutpoint falling into an intermediary zone. Within a given cohort, reference ranges and corresponding intermediary zones can help facilitate the development of a research-oriented system that would require fewer confirmatory Aβ-PET scans, freeing up potential resources that can be allocated toward acquiring tau-PET or other ADRD biomarkers [8]. As noted above, combining optimal (Youden index = .338 pg/mL) and two-point ROC detection thresholds (max-sensitivity = .253; max-specificity = .472) stratified the cohort into four zones that best defined amyloid positivity at *baseline*, and captured the heterogeneity in our cohort, and further helped minimize and refine an intermediary zone. In the current study, exploratory stratification analyses were provided in the supplemental materials to permit general comparisons across studies and to provide a detailed account of how demographic and health factors contribute to the cutpoints and range of thresholds we observed using the entire cohort. This descriptive approach is meant to provide a foundation for assessing the extent to which cutpoints and reference ranges are stable over time and sensitive to amyloid accumulation. We of course recognize that model-based approaches are preferred when cutpoints are to be used for clinical decision-making, and to predict dementia risk, to circumvent additional sampling biases.

A strength of the study is the comprehensive assessment of demographics and chronic conditions on cutpoints and discordance, as well as the consideration of the time interval between the blood draw and PET scan. Although the interval between blood collection was not associated with biomarker levels nor impact any of the GLMs, restricting analyses to scans within 14 months of blood draw provided the greatest discrimination of Aβ-PET-from Aβ-PET+ participants across amyloid PET thresholds. In exploratory analyses, we observed an interesting pattern such that participants with higher baseline levels of p-tau217 who were Aβ-PET negative (p-tau217+/AB-PET-) were more likely to have a history of hypertension, whereas the few participants with low p-tau217 levels considered Aβ-PET positive (p-tau217-/AB-PET+) had poorer cardiometabolic health, were Black, and female. Further, p-tau217 levels and cutpoints were slightly lower, on average, in cognitively unimpaired participants and those who self-identified as female or as Black or African American, and was as in *APOE*-ε4 non-carriers. However, the number of discordant cases were relatively small, and validation of this finding is needed. However, limitations also warrant consideration. This study was cross-sectional. Processing of longitudinal blood samples is ongoing and will allow for a long-term assessment of p-tau217 cutpoints and reference ranges and individual trajectories in our cohort.

## CONCLUSION

As previously reported, p-tau217 is a highly specific marker of cerebral amyloid burden. When evaluating the utility of p-tau217 to robustly capture Aβ-PET deposition, studies should minimize the time elapsed between blood sample collection and amyloid-PET scan acquisition. In addition to considering how sex, race, APOE *jointly* interact to convey risk for amyloid pathology and dementia, common comorbidities and health risk factors such as BMI and kidney function and cardiometabolic health should be evaluated carefully, particularly when monitoring intermediate (positive and negative) cases. Additional longitudinal analyses assessing how vascular and metabolic factors may impact disease progression captured by p-tau217 are needed, particularly in more diverse and heterogeneous cohorts [7,59–61].

## Consent Statement

Written informed consent was obtained for all participants and/or their legally authorized representatives.

## Data Availability

All data produced in the present study are available upon reasonable request to the authors

## Acknowledgements & Funding

This work was supported by the Wake Forest University School of Medicine’s Alzheimer’s Disease Research Center (P30AG049638, P30AG072947, R01AG054069, R01AG058969, and T32AG033534), which is funded by the National Institute on Aging (NIA). Additional support was provided by the Department of Gerontology and Geriatric Medicine and Center for Healthy Aging and Alzheimer’s Prevention, Wake Forest School of Medicine. Plasma biomarker analyses were completed in part by the NCRAD Biomarker Assay Laboratory as part of the Alzheimer’s Disease Center Fluid Biomarker (ADCFB) Initiative, which receives government support under a cooperative agreement grant (U24 AG021886) awarded by the National Institute on Aging (NIA). Plasma p-tau217 analytes were processed at Neurocode (Bellingham, WA) and funded by ALZpath (Carlsbad CA). This study would not have been possible without the commitment and support of our valued WFADRC staff and study participants.

This study was supported by the following funding sources: Rudolph reports funding for this work from National Institutes of Health (NIH) P30AG072947 and T32AG033534. Sutphen reports funding for this work from NIH P30AG049638 and P30AG072947 and additional funding from other NIH grants to the institution, including T32AG033534. Register reports funding for this work from NIH P30AG049638 and P30AG072947 and additional funding from other NIH grants to the institution. Lockhart reports funding for this work from NIH P30AG072947 and additional funding from other NIH grants to the institution. Rundle reports funding for this work from NIH P30AG072947 and additional funding from other NIH grants to the institution. Hughes reports funding for this work from NIH P30AG072947 and additional funding from other NIH grants to the institution. Bateman reports funding for this work from NIH P30AG072947, other NIH grants, and funding from ASPECT 20-AVP-786-306 to the institution. Solingapuram Sai reports funding for this work from NIH P30AG072947 and additional funding from other NIH grants to the institution. Whitlow reports funding for this work from NIH P30AG072947 and additional funding from other NIH grants to the institution. Craft reports funding for this work from NIH P30AG072947 and additional funding from other NIH grants. Mielke reports funding for this work from NIH P30AG072947 and U24 AG082930.

## REFERENCES

[1] Rudolph MD, Sutphen CL, Register TC, Whitlow CT, Solingapuram Sai KK, Hughes TM, et al. Associations among plasma, MRI, and amyloid PET biomarkers of Alzheimer’s disease and related dementias and the impact of health-related comorbidities in a community-dwelling cohort. Alzheimer’s & Dementia 2024;20:4159. 10.1002/ALZ.13835.

[2] Hansson O, Edelmayer RM, Boxer AL, Carrillo MC, Mielke MM, Rabinovici GD, et al. The Alzheimer’s Association appropriate use recommendations for blood biomarkers in Alzheimer’s disease. Alzheimer’s and Dementia 2022;18:2669–86. 10.1002/alz.12756.

[3] Teunissen CE, Verberk IMW, Thijssen EH, Vermunt L, Hansson O, Zetterberg H, et al. Blood-based biomarkers for Alzheimer’s disease: towards clinical implementation. Lancet Neurol 2022;21:66–77. 10.1016/S1474-4422(21)00361-6.

[4] Jack CR, Wiste HJ, Algeciras-Schimnich A, Figdore DJ, Schwarz CG, Lowe VJ, et al. Predicting amyloid PET and tau PET stages with plasma biomarkers. Brain 2023;146:2029–44. 10.1093/BRAIN/AWAD042.

[5] Alcolea D, Delaby C, Muñoz L, Torres S, Estellés T, Zhu N, et al. Use of plasma biomarkers for AT(N) classification of neurodegenerative dementias. J Neurol Neurosurg Psychiatry 2021;92. 10.1136/jnnp-2021-326603.

[6] Palmqvist S, Stomrud E, Cullen N, Janelidze S, Manuilova E, Jethwa A, et al. An accurate fully automated panel of plasma biomarkers for Alzheimer’s disease. Alzheimer’s & Dementia 2023;19:1204–15. 10.1002/ALZ.12751.

[7] Mielke MM, Dage JL, Frank RD, Algeciras-Schimnich A, Knopman DS, Lowe VJ, et al. Performance of plasma phosphorylated tau 181 and 217 in the community. Nature Medicine 2022 28:7 2022;28:1398–405. 10.1038/s41591-022-01822-2.

[8] Mattsson-Carlgren N, Collij LE, Stomrud E, Pichet Binette A, Ossenkoppele R, Smith R, et al. Plasma Biomarker Strategy for Selecting Patients With Alzheimer Disease for Antiamyloid Immunotherapies. JAMA Neurol 2023. 10.1001/JAMANEUROL.2023.4596.

[9] Benedet AL, Brum WS, Hansson O, Karikari TK, Zimmer ER, Zetterberg H, et al. The accuracy and robustness of plasma biomarker models for amyloid PET positivity. Alzheimers Res Ther 2022;14:1–11. 10.1186/s13195-021-00942-0.

[10] Ashton NJ, Wagner ;, Brum S, Guglielmo;, Molfetta D, Benedet AL, et al. Diagnostic Accuracy of a Plasma Phosphorylated Tau 217 Immunoassay for Alzheimer Disease Pathology. JAMA Neurol 2024. 10.1001/JAMANEUROL.2023.5319.

[11] Thijssen EH, La Joie R, Wolf A, Strom A, Wang P, Iaccarino L, et al. Diagnostic value of plasma phosphorylated tau181 in Alzheimer’s disease and frontotemporal lobar degeneration. Nat Med 2020;26:387–97. 10.1038/s41591-020-0762-2.

[12] Palmqvist S, Janelidze S, Quiroz YT, Zetterberg H, Lopera F, Stomrud E, et al. Discriminative Accuracy of Plasma Phospho-tau217 for Alzheimer Disease vs Other Neurodegenerative Disorders. JAMA 2020;324:772–81. 10.1001/JAMA.2020.12134.

[13] Schindler SE, Galasko D, Pereira AC, Rabinovici GD, Salloway S, Suárez-Calvet M, et al. Acceptable performance of blood biomarker tests of amyloid pathology — recommendations from the Global CEO Initiative on Alzheimer’s Disease. Nat Rev Neurol 2024;20:426–39. 10.1038/S41582-024-00977-5.

[14] Schindler SE, Petersen KK, Saef B, Tosun D, Shaw LM, Zetterberg H, et al. Head-to-head comparison of leading blood tests for Alzheimer’s disease pathology. Alzheimer’s & Dementia 2024;20:8074. 10.1002/ALZ.14315.

[15] Milà-Alomà M, Ashton NJ, Shekari M, Salvadó G, Ortiz-Romero P, Montoliu-Gaya L, et al. Plasma p-tau231 and p-tau217 as state markers of amyloid-β pathology in preclinical Alzheimer’s disease. Nature Medicine 2022 28:9 2022;28:1797–801. 10.1038/s41591-022-01925-w.

[16] Janelidze S, Bali D, Ashton NJ, Barthélemy NR, Vanbrabant J, Stoops E, et al. Head-to-head comparison of 10 plasma phospho-tau assays in prodromal Alzheimer’s disease. Brain 2023;146:1592–601. 10.1093/BRAIN/AWAC333.

[17] Lehmann S, Schraen-Maschke S, Vidal JS, Delaby C, Buee L, Blanc F, et al. Clinical value of plasma ALZpath pTau217 immunoassay for assessing mild cognitive impairment. J Neurol Neurosurg Psychiatry 2024;0:1–8. 10.1136/JNNP-2024-333467.

[18] Syrjanen JA, Campbell MR, Algeciras-Schimnich A, Vemuri P, Graff-Radford J, Machulda MM, et al. Associations of amyloid and neurodegeneration plasma biomarkers with comorbidities. Alzheimer’s and Dementia 2022;18:1128–40. 10.1002/alz.12466.

[19] Mielke MM, Fowler NR. Alzheimer disease blood biomarkers: considerations for population-level use. Nature Reviews Neurology 2024 20:8 2024;20:495–504. 10.1038/s41582-024-00989-1.

[20] Westwood S, Liu B, Baird AL, Anand S, Nevado-Holgado AJ, Newby D, et al. The influence of insulin resistance on cerebrospinal fluid and plasma biomarkers of Alzheimer’s pathology. Alzheimers Res Ther 2017;9. 10.1186/s13195-017-0258-6.

[21] van Arendonk J, Neitzel J, Steketee RME, van Assema DME, Vrooman HA, Segbers M, et al. Diabetes and hypertension are related to amyloid-beta burden in the population-based Rotterdam Study. Brain 2023;146:337. 10.1093/BRAIN/AWAC354.

[22] Gottesman RF, Schneider ALC, Zhou Y, Coresh J, Green E, Gupta N, et al. Association between midlife vascular risk factors and estimated brain amyloid deposition. JAMA 2017;317:1443. 10.1001/JAMA.2017.3090.

[23] Keuss SE, Coath W, Nicholas JM, Poole T, Barnes J, Cash DM, et al. Associations of β-Amyloid and Vascular Burden With Rates of Neurodegeneration in Cognitively Normal Members of the 1946 British Birth Cohort. Neurology 2022;99:E129–41. 10.1212/WNL.0000000000200524.

[24] Coffin C, Suerken CK, Bateman JR, Whitlow CT, Williams BJ, Espeland MA, et al. Vascular and microstructural markers of cognitive pathology. Alzheimer’s & Dementia : Diagnosis, Assessment & Disease Monitoring 2022;14. 10.1002/DAD2.12332.

[25] Albert MS, DeKosky ST, Dickson D, Dubois B, Feldman HH, Fox NC, et al. The diagnosis of mild cognitive impairment due to Alzheimer’s disease: Recommendations from the National Institute on Aging-Alzheimer’s Association workgroups on diagnostic guidelines for Alzheimer’s disease. Alzheimers Dement 2011;7:270. 10.1016/J.JALZ.2011.03.008.

[26] McKhann GM, Knopman DS, Chertkow H, Hyman BT, Jack CR, Kawas CH, et al. The diagnosis of dementia due to Alzheimer’s disease: Recommendations from the National Institute on Aging-Alzheimer’s Association workgroups on diagnostic guidelines for Alzheimer’s disease. Alzheimers Dement 2011;7:263. 10.1016/J.JALZ.2011.03.005.

[27] Hughes TM, Lockhart SN, Suerken CK, Jung Y, Whitlow CT, Bateman JR, et al. Hypertensive Aspects of Cardiometabolic Disorders Are Associated with Lower Brain Microstructure, Perfusion, and Cognition. J Alzheimers Dis 2022;90:1589. 10.3233/JAD-220646.

[28] Wakabayashi I, Daimon T. The “cardiometabolic index” as a new marker determined by adiposity and blood lipids for discrimination of diabetes mellitus. Clinica Chimica Acta 2015;438:274–8. 10.1016/J.CCA.2014.08.042.

[29] Weintraub S, Besser L, Dodge HH, Teylan M, Ferris S, Goldstein FC, et al. Version 3 of the Alzheimer Disease Centers’ Neuropsychological Test Battery in the Uniform Data Set (UDS). Alzheimer Dis Assoc Disord 2018;32:10–7. 10.1097/WAD.0000000000000223.

[30] Lockhart SN, Schaich CL, Craft S, Sachs BC, Rapp SR, Jung Y, et al. Associations among vascular risk factors, neuroimaging biomarkers, and cognition: Preliminary analyses from the Multi-Ethnic Study of Atherosclerosis (MESA). Alzheimer’s & Dementia 2022;18:551–60. 10.1002/ALZ.12429.

[31] Klunk WE, Engler H, Nordberg A, Wang Y, Blomqvist G, Holt DP, et al. Imaging brain amyloid in Alzheimer’s disease with Pittsburgh Compound-B. Ann Neurol 2004;55:306–19. 10.1002/ANA.20009.

[32] Mormino EC, Brandel MG, Madison CM, Rabinovici GD, Marks S, Baker SL, et al. Not quite PIB-positive, not quite PIB-negative: slight PIB elevations in elderly normal control subjects are biologically relevant. Neuroimage 2012;59:1152. 10.1016/J.NEUROIMAGE.2011.07.098.

[33] Klunk WE, Koeppe RA, Price JC, Benzinger TL, Devous MD, Jagust WJ, et al. The Centiloid Project: Standardizing quantitative amyloid plaque estimation by PET. Alzheimer’s & Dementia 2015;11:1–15.e4. 10.1016/J.JALZ.2014.07.003.

[34] Buckley CJ, Sherwin PF, Smith APL, Wolber J, Weick SM, Brooks DJ. Validation of an electronic image reader training programme for interpretation of [18F]flutemetamol β-amyloid PET brain images. Nucl Med Commun 2016;38:234. 10.1097/MNM.0000000000000633.

[35] Collij LE, Salvadó G, Shekari M, Lopes Alves I, Reimand J, Wink AM, et al. Visual assessment of [18F]flutemetamol PET images can detect early amyloid pathology and grade its extent. Eur J Nucl Med Mol Imaging 2021;48:2169–82. 10.1007/S00259-020-05174-2/FIGURES/6.

[36] Villeneuve S, Rabinovici GD, Cohn-Sheehy BI, Madison C, Ayakta N, Ghosh PM, et al. Existing Pittsburgh Compound-B positron emission tomography thresholds are too high: statistical and pathological evaluation. Brain 2015;138:2020. 10.1093/BRAIN/AWV112.

[37] Knopman DS, Lundt ES, Therneau TM, Albertson SM, Gunter JL, Senjem ML, et al. Association of Initial β-Amyloid Levels With Subsequent Flortaucipir Positron Emission Tomography Changes in Persons Without Cognitive Impairment. JAMA Neurol 2021;78:1. 10.1001/JAMANEUROL.2020.3921.

[38] La Joie R, Ayakta N, Seeley WW, Borys E, Boxer AL, DeCarli C, et al. Multi-site study of the relationships between ante mortem [11C]PIB-PET Centiloid values and post mortem measures of Alzheimer’s disease neuropathology. Alzheimers Dement 2019;15:205. 10.1016/J.JALZ.2018.09.001.

[39] Amft M, Ortner M, Eichenlaub U, Goldhardt O, Diehl-Schmid J, Hedderich DM, et al. The cerebrospinal fluid biomarker ratio Aβ42/40 identifies amyloid positron emission tomography positivity better than Aβ42 alone in a heterogeneous memory clinic cohort. Alzheimers Res Ther 2022;14. 10.1186/S13195-022-01003-W.

[40] Pérez-Grijalba V, Romero J, Pesini P, Sarasa L, Monleón I, San-José I, et al. Plasma Aβ42/40 Ratio Detects Early Stages of Alzheimer’s Disease and Correlates with CSF and Neuroimaging Biomarkers in the AB255 Study. J Prev Alzheimers Dis 2019;6:34–41. 10.14283/JPAD.2018.41.

[41] Salvadó G, Molinuevo JL, Brugulat-Serrat A, Falcon C, Grau-Rivera O, Suárez-Calvet M, et al. Centiloid cut-off values for optimal agreement between PET and CSF core AD biomarkers. Alzheimers Res Ther 2019;11. 10.1186/S13195-019-0478-Z.

[42] Sutphen CL, Rudolph MD, Lockhart SN, Whitlow CT, Solingapuram Sai KK, Hughes TM, et al. Comparison of plasma Ab42, Ab40, and p-tau181 assessed on two different platforms with CSF AD biomarkers and amyloid (PiB) PET. Alzheimer’s & Dementia 2023;19:e079859. 10.1002/ALZ.079859.

[43] Farrell ME, Jiang S, Schultz AP, Properzi MJ, Price JC, Becker JA, et al. Defining the Lowest Threshold for Amyloid-PET to Predict Future Cognitive Decline and Amyloid Accumulation. Neurology 2021;96:e619–31. 10.1212/WNL.0000000000011214.

[44] Jansen WJ, Janssen O, Tijms BM, Vos SJB, Ossenkoppele R, Visser PJ, et al. Prevalence Estimates of Amyloid Abnormality Across the Alzheimer Disease Clinical Spectrum. JAMA Neurol 2022;79:228–43. 10.1001/JAMANEUROL.2021.5216.

[45] Benaglia T, Chauveau D, Hunter DR, Young DS. mixtools: An R Package for Analyzing Mixture Models. J Stat Softw 2010;32:1–29. 10.18637/JSS.V032.I06.

[46] Thiele C, Hirschfeld G. cutpointr: Improved Estimation and Validation of Optimal Cutpoints in R. J Stat Softw 2021;98:1–27. 10.18637/JSS.V098.I11.

[47] Robin X, Turck N, Hainard A, Tiberti N, Lisacek F, Sanchez JC, et al. pROC: An open-source package for R and S+ to analyze and compare ROC curves. BMC Bioinformatics 2011;12:1–8. 10.1186/1471-2105-12-77/TABLES/3.

[48] Kassambra A. Package “rstatix”: Pipe-Friendly Framework for Basic Statistical Tests 2023.

[49] Benjamini Y, Hochberg Y. Controlling the false discovery rate: a practical and powerful approach to multiple testing. Journal of the Royal Statistical Society 1995;57:289–300. 10.2307/2346101.

[50] Delanaye P, Mariat C. The applicability of eGFR equations to different populations. Nat Rev Nephrol 2013;9:513–22. 10.1038/NRNEPH.2013.143.

[51] Stevens LA, Levey AS. Measured GFR as a confirmatory test for estimated GFR. J Am Soc Nephrol 2009;20:2305–13. 10.1681/ASN.2009020171.

[52] Ankeny SE, Bacci JR, Decourt B, Sabbagh MN, Mielke MM, Ankeny SE, et al. Navigating the Landscape of Plasma Biomarkers in Alzheimer’s Disease: Focus on Past, Present, and Future Clinical Applications. Neurology and Therapy 2024 2024:1–17. 10.1007/S40120-024-00658-X.

[53] della Monica C, Revell V, Atzori G, Laban R, Skene SS, Heslegrave A, et al. P-tau217 and other blood biomarkers of dementia: variation with time of day. Translational Psychiatry 2024 14:1 2024;14:1–9. 10.1038/s41398-024-03084-7.

[54] Giangrande C, Delatour V, Andreasson U, Blennow K, Gobom J, Zetterberg H. Harmonization and standardization of biofluid-based biomarker measurements for AT(N) classification in Alzheimer’s disease. Alzheimer’s and Dementia: Diagnosis, Assessment and Disease Monitoring 2023;15. 10.1002/dad2.12465.

[55] Hansson O, Blennow K, Zetterberg H, Dage J. Blood biomarkers for Alzheimer’s disease in clinical practice and trials. Nat Aging 2023;3:506–19. 10.1038/S43587-023-00403-3.

[56] O’Bryant SE, Petersen M, Hall J, Johnson LA. Medical comorbidities and ethnicity impact plasma Alzheimer’s disease biomarkers: Important considerations for clinical trials and practice. Alzheimers Dement 2023;19:36–43. 10.1002/ALZ.12647.

[57] Rowe CC, Pejoska S, Mulligan RS, Jones G, Chan JG, Svensson S, et al. Head-to-Head Comparison of 11C-PiB and 18F-AZD4694 (NAV4694) for β-Amyloid Imaging in Aging and Dementia. Journal of Nuclear Medicine 2013;54:880–6. 10.2967/JNUMED.112.114785.

[58] Lowe VJ, Mester CT, Lundt ES, Lee J, Ghatamaneni S, Algeciras-Schimnich A, et al. Amyloid PET detects the deposition of brain Aβ earlier than CSF fluid biomarkers. Alzheimer’s & Dementia 2024;20:8097–112. 10.1002/ALZ.14317.

[59] Hampel H, Hu Y, Cummings J, Mattke S, Iwatsubo T, Nakamura A, et al. Blood-based biomarkers for Alzheimer’s disease: Current state and future use in a transformed global healthcare landscape. Neuron 2023;111:2781–99. 10.1016/J.NEURON.2023.05.017/ASSET/2042E132-9024-4A16-ACCD-5476B15AA238/MAIN.ASSETS/GR1.JPG.

[60] Schindler SE, Karikari TK, Ashton NJ, Henson RL, Yarasheski KE, West T, et al. Effect of Race on Prediction of Brain Amyloidosis by Plasma Aβ42/Aβ40, Phosphorylated Tau, and Neurofilament Light. Neurology 2022;99:E245–57. 10.1212/WNL.0000000000200358.

[61] Pichet Binette A, Janelidze S, Cullen N, Dage JL, Bateman RJ, Zetterberg H, et al. Confounding factors of Alzheimer’s disease plasma biomarkers and their impact on clinical performance. Alzheimer’s and Dementia 2022. 10.1002/alz.12787.

