## Supplemental Data for "Evaluation of plasma p-tau217 for detecting amyloid pathology in a diverse and heterogeneous community-based cohort"

### SUPPLEMENTARY MATERIALS

#### 1 SUPPLEMENTARY RESULTS

##### Discordance and medical conditions

We did not exclude participants based on history of stroke (UDS: CBSTROKE), traumatic brain injury (UDS: TBI), or myocardial infarction (UDS: MYOINF). Evaluating the impact of these health conditions, we did not observe any discordant participants (*n*=38) who reported a history of stroke. A single participant, p-tau217-/AB-PET+ at baseline, reported having a recent/active history of TBI. 4 of 38 discordant participants had a recent/active history of myocardial infarction (3 p-tau+/Aβ-PET-; 1 p-tau-/Aβ-PET+ at baseline).

#### 2 SUPPLEMENTARY TABLES

##### Table S1. Participant characteristics by amyloid PET positivity status (CL ≥ 24)

| **Characteristic** | **Overall (*n*=307)[1]** | **Aβ-PET- (*n*=202)[1]** | **Aβ-PET+ (n=105)[1]** | ***p*[2]** |
|---|---|---|---|---|
| **Dx** | | | | <0.001 |
| CU | 162 (53%) | 132 (65%) | 30 (29%) | |
| MCI | 106 (35%) | 59 (29%) | 47 (45%) | |
| DEM | 35 (11%) | 9 (4.5%) | 26 (25%) | |
| OTHER | 4 (1.3%) | 2 (1.0%) | 2 (1.9%) | |
| **Age** | **69.34 (7.80)** | **67.70 (7.33)** | **72.49 (7.74)** | **<0.001** |
| **Female** | 182 (59%) | 119 (59%) | 63 (60%) | 0.9 |
| **Race** | | | | 0.12 |
| American Indian or Alaska Native | 2 (0.7%) | 1 (0.5%) | 1 (1.0%) | |
| Asian | 3 (1.0%) | 3 (1.5%) | 0 (0%) | |
| Black or African American | 42 (14%) | 33 (16%) | 9 (8.6%) | |
| White | 260 (85%) | 165 (82%) | 95 (90%) | |
| **Education (Years)** | **16.04 (2.58)** | **16.28 (2.45)** | **15.58 (2.75)** | **0.035** |
| ***APOE*-ε4 Carriers** | **91 (30%)** | **41 (21%)** | **50 (50%)** | **<0.001** |
| **eGFR** | **79.09 (14.01)** | **80.61 (13.42)** | **76.15 (14.71)** | **0.022** |
| **BMI** | **27.56 (5.34)** | **28.05 (5.67)** | **26.61 (4.52)** | **0.042** |
| Glucose | 103.88 (23.48) | 104.41 (23.80) | 102.87 (22.97) | 0.3 |
| Hemoglobin A1c | 5.68 (0.47) | 5.69 (0.53) | 5.64 (0.34) | 0.7 |
| LDL Cholesterol | 99.01 (31.14) | 98.47 (29.58) | 100.06 (34.32) | >0.9 |
| HDL Cholesterol | 62.07 (21.31) | 61.85 (22.03) | 62.49 (19.99) | 0.7 |
| OGTT (Baseline) | 95.10 (15.43) | 95.41 (16.22) | 94.53 (13.89) | 0.5 |
| OGTT (120) | 136.23 (43.27) | 133.53 (42.12) | 142.03 (45.43) | 0.2 |
| CMI | -0.85 (0.52) | -0.88 (0.55) | -0.79 (0.47) | 0.053 |
| Diabetes Group | | | | >0.9 |
| Diabetes | 27 (8.9%) | 18 (9.0%) | 9 (8.7%) | |
| Normal | 130 (43%) | 87 (44%) | 43 (41%) | |
| Prediabetes | 147 (48%) | 95 (48%) | 52 (50%) | |
| HTN HX | 119 (39%) | 73 (36%) | 46 (44%) | 0.2 |
| **p-tau181** | **3.38 (1.86)** | **2.82 (1.21)** | **4.49 (2.35)** | **<0.001** |
| **p-tau217** | **0.43 (0.35)** | **0.26 (0.14)** | **0.76 (0.39)** | **<0.001** |
| **SUVr** | **1.39 (0.44)** | **1.11 (0.08)** | **1.92 (0.36)** | **<0.001** |

| | | | | |
|---|---|---|---|---|
| **Centiloids** | **27.11 (43.77)** | **-0.62 (7.16)** | **80.47 (34.21)** | **<0.001** |
| Blood-PET Interval* | -415.18 (580.68) | -423.32 (586.43) | -399.53 (571.92) | 0.8 |

[1] n (%); Mean (SD)

[2] Fisher's exact test; Wilcoxon rank sum test; Pearson's Chi-squared test

*Time interval between blood collection and amyloid PET scan

#### Table S2. Optimal and two-point detection thresholds for p-tau217

| | | | | | | | | | | | | **TWO-POINT DETECTION THRESHOLD** | | | | | | | | | |
|---|---|---|---|---|---|---|---|---|---|---|---|---|---|---|---|---|---|---|---|---|---|
| | | **Optimal Cutpoints (Youden Index)** | | | | | | | | | | **MAX-SENS (LOWER BOUND)** | | | | | **MAX-SPEC (UPPER BOUND)** | | | | |
| **n** | **Aβ-PET+** | **prev** | **cp** | **bootCP** | **auc** | **bootAUC** | **acc** | **sens** | **spec** | **ppv** | **npv** | **thresh** | **sens** | **spec** | **ppv** | **npv** | **thresh** | **sens** | **spec** | **ppv** | **npv** |
| 316 | VISREAD | .364 | **.339** | .302-.412 | .955 | .929-.976 | .899 | .923 | .885 | .821 | .953 | **.303** | .952 | .813 | .744 | .967 | **.429** | .808 | .951 | .903 | .896 |
| | SUVR≥1.21 | .412 | **.338** | .302-.406 | .914 | .880-.945 | .863 | .829 | .886 | .836 | .881 | **.232** | .953 | .549 | .597 | .944 | **.417** | .705 | .951 | .910 | .822 |
| | CL≥12 | .398 | **.338** | .302-.404 | .926 | .893-.955 | .875 | .851 | .891 | .837 | .901 | **.240** | .950 | .596 | .608 | .948 | **.460** | .711 | .951 | .905 | .833 |
| 307 | CL≥24 | .342 | **.338** | .322-.460 | .942 | .914-.966 | .878 | .914 | .856 | .768 | .951 | **.253** | .952 | .624 | .568 | .962 | **.472** | .743 | .950 | .886 | .878 |
| | CL≥32 | .316 | **.341** | .341-.465 | .956 | .931-.976 | .882 | .948 | .851 | .746 | .973 | **.340** | .948 | .851 | .746 | .973 | **.483** | .792 | .952 | .884 | .908 |
| | | | | | | | | | | | | **CU Only** | | | | | | | | | |
| **n** | **Aβ-PET+** | **prev** | **cp** | **bootCP** | **auc** | **bootAUC** | **acc** | **sens** | **spec** | **ppv** | **npv** | **thresh** | **sens** | **spec** | **ppv** | **npv** | **thresh** | **sens** | **spec** | **ppv** | **npv** |
| 169 | VISREAD | .194 | **.341** | .305-.371 | .915 | .858-.961 | .877 | .833 | .888 | .641 | .957 | **.247** | .933 | .608 | .364 | .974 | **.399** | .667 | .952 | .769 | .922 |
| | SUVR≥1.21 | .235 | **.338** | .231-.371 | .839 | .758-.909 | .843 | .692 | .890 | .659 | .904 | **.194** | .949 | .346 | .308 | .957 | **.399** | .538 | .953 | .778 | .871 |
| | CL≥12 | .226 | **.302** | .231-.404 | .858 | .782-.921 | .805 | .778 | .813 | .549 | .926 | **.229** | .944 | .561 | .386 | .972 | **.399** | .556 | .951 | .769 | .880 |
| 162 | CL≥24 | .185 | **.338** | .234-.371 | .898 | .833-.949 | .868 | .800 | .879 | .600 | .951 | **.242** | .933 | .591 | .341 | .975 | **.405** | .600 | .946 | .720 | .912 |
| | CL≥32 | .164 | **.341** | .305-.406 | .925 | .870-.969 | .881 | .846 | .887 | .595 | .967 | **.247** | .962 | .609 | .325 | .988 | **.405** | .692 | .947 | .720 | .940 |
| | | | | | | | | | | | | **Blood-PET Interval < 14 months** | | | | | | | | | |
| n | Aβ-PET+ | **prev** | **cp** | **bootCP** | **auc** | **bootAUC** | **acc** | **sens** | **spec** | **ppv** | **npv** | **thresh** | **sens** | **spec** | **ppv** | **npv** | **thresh** | **sens** | **spec** | **ppv** | **npv** |
| 213 | VISREAD | .374 | **.351** | .322-.460 | .972 | .948-.990 | .924 | .959 | .903 | .855 | .974 | **.357** | .946 | .911 | .864 | .966 | **.460** | .851 | .952 | .913 | .915 |
| | SUVR≥1.21 | .418 | **.338** | .303-.370 | .934 | .895-.967 | .892 | .899 | .887 | .851 | .924 | **.269** | .955 | .685 | .685 | .955 | **.460** | .742 | .952 | .917 | .837 |
| | CL≥12 | .410 | **.338** | .316-.370 | .957 | .924-.983 | .915 | .931 | .904 | .871 | .950 | **.316** | .954 | .864 | .830 | .964 | **.397** | .816 | .952 | .922 | .881 |
| 214 | CL≥24 | .363 | **.338** | .322-.460 | .967 | .943-.986 | .906 | .974 | .867 | .798 | .983 | **.348** | .948 | .882 | .820 | .968 | **.460** | .831 | .949 | .901 | .908 |
| | CL≥32 | .349 | **.351** | .322-.461 | .967 | .944-.986 | .901 | .959 | .870 | .855 | .974 | **.348** | .948 | .881 | .820 | .967 | **.460** | .831 | .948 | .901 | .908 |
| | | | | | | | | | | | | **CU Only** | | | | | | | | | |
| **n** | **Aβ-PET+** | **prev** | **cp** | **bootCP** | **auc** | **bootAUC** | **acc** | **sens** | **spec** | **ppv** | **npv** | **thresh** | **sens** | **spec** | **ppv** | **npv** | **thresh** | **sens** | **spec** | **ppv** | **npv** |
| | VISREAD | .152 | **.305** | .305-.351 | .938 | .882-.981 | .826 | 1.00 | .795 | .484 | .969 | **.193** | .941 | .321 | .232 | .962 | **.366** | .588 | .949 | .714 | .914 |
| | SUVR≥1.21 | .179 | **.305** | .305-.351 | .863 | .741-.958 | .811 | .882 | .795 | .467 | 1.00 | **.319** | .929 | .846 | .520 | .985 | **.366** | .714 | .949 | .714 | .949 |
| 96 | CL≥12 | .168 | **.305** | .305-.371 | .898 | .798-.969 | .821 | .938 | .797 | .484 | .984 | **.303** | .938 | .797 | .484 | .984 | **.366** | .625 | .949 | .714 | .926 |
| | CL≥24 | .156 | **.305** | .305-.371 | .934 | .881-.979 | .832 | 1.00 | .790 | .484 | 1.00 | **.319** | .933 | .850 | .538 | .985 | **.366** | .667 | .950 | .714 | .938 |
| | CL≥32 | .147 | **.305** | .305-.351 | .938 | .884-.982 | .821 | 1.00 | .790 | .798 | .976 | **.357** | .946 | .877 | .805 | .968 | **.485** | .797 | .949 | .894 | .897 |

*Note:* Two-point detection thresholds maximizing sensitivity (SENS) or specificity (SPEC) before and after excluding large intervals between blood collection and scan acquisition (< 14 months) and restricting the sample to cognitively normal participants. Abbreviations: thresh= threshold (lower or upper boundary); sens = sensitivity; spec = specificity; ppv = positive predictive value; npv = negative predictive values.

**Table S3. Participant characteristics by zone based on 4-tier system combining two-point detection and binary p-tau217 cutpoints**

| | N | Overall N = 598[1] | NEG (< .253) n = 241[1] | INT-LOW (.253-.338) n = 109[1] | INT-HIGH (.338-.472) n = 77[1] | POS (≥ .472) n = 171[1] |
|---|---|---|---|---|---|---|
| Dx | 598 | | | | | |
| CU | | 314 (53%) | 156 (65%) | 68 (62%) | 45 (58%) | 45 (26%) |
| MCI | | 213 (36%) | 73 (30%) | 36 (33%) | 28 (36%) | 76 (44%) |
| DEM | | 64 (11%) | 9 (3.7%) | 4 (3.7%) | 2 (2.6%) | 49 (29%) |
| OTHER | | 7 (1.2%) | 3 (1.2%) | 1 (0.9%) | 2 (2.6%) | 1 (0.6%) |
| Age (Baseline) | 598 | 69 (63, 76) | 66 (62, 72) | 70 (64, 75) | 71 (66, 76) | 74 (68, 79) |
| Education | 598 | 16 (14, 18) | 16 (14, 18) | 16 (14, 18) | 16 (14, 18) | 16.00 (13, 18) |
| Female | 598 | 388 (65%) | 172 (71%) | 69 (63%) | 47 (61%) | 100 (58%) |
| Race | 598 | 124 (21%) | 72 (30%) | 19 (17%) | 11 (14%) | 22 (13%) |
| *APOE*-ε4 Carriers | 579 | 187 (32%) | 55 (23%) | 28 (26%) | 23 (31%) | 81 (50%) |
| eGFR | 552 | 79 (68, 90) | 82 (72, 92) | 79 (69, 88) | 77 (69, 88) | 74 (65, 86) |
| BMI | 598 | 26.8 (24.1, 30.9) | 27.7 (24.5, 32.0) | 27.1 (24.1, 31.5) | 26.5 (24.7, 30.3) | 25.7 (22.8, 29.4) |
| CMI | 587 | -0.84 (-1.16, -0.51) | -0.86 (-1.17, -0.47) | -0.97 (-1.20, -0.64) | -0.78 (-1.13, -0.44) | -0.77 (-1.12, -0.53) |
| Insulin | 484 | 13 (8, 27) | 14 (8, 31) | 14 (7, 34) | 12 (9, 24) | 12 (8, 23) |
| Glucose | 413 | 98 (90, 110) | 98 (91, 111) | 97 (91, 107) | 101 (92, 110) | 98 (89, 110) |
| HTN HX | 598 | 260 (43%) | 100 (41%) | 46 (42%) | 31 (40%) | 83 (49%) |
| HTN MEDS | 598 | 283 (47%) | 112 (46%) | 45 (41%) | 34 (44%) | 92 (54%) |
| ANY MEDS | 598 | 579 (97%) | 232 (96%) | 106 (97%) | 74 (96%) | 167 (98%) |
| p-tau217 | 598 | 0.30 (0.21, 0.53) | 0.19 (0.16, 0.22) | 0.30 (0.28, 0.31) | 0.39 (0.36, 0.43) | 0.78 (0.60, 1.12) |
| AB-PET (SUVr) | 316 | 1.15 (1.07, 1.65) | 1.08 (1.05, 1.13) | 1.12 (1.06, 1.18) | 1.39 (1.13, 1.63) | 1.97 (1.67, 2.21) |
| AB-PET (CL) | 307 | 2 (-4, 55) | -3 (-6, 1) | -1 (-5, 6) | 22 (0, 51) | 90 (56, 109) |
| Blood- PET Interval | 330 | -125 (-731, -36) | -123 (-946, -27) | -140 (-489, -37) | -141 (-594, -40) | -109 (-402, -43) |

[1] n (%); Median (IQR); [2] Pearson's Chi-squared test; [3] False discovery rate correction

*Note*: Primary cutpoints established using a CL ≥ 24 to define amyloid PET positivity.

**Table S4. Discordance across plasma p-tau217 cutpoints before and after excluding participants with intervals greater than 14 months between blood collection and scan acquisition**

A

| | Amyloid (Aβ)-PET POSITIVITY | | | | | ROC ANALYSES | |
|---|---|---|---|---|---|---|---|
| **Discordance** | **VISUAL READ** | **SUVR ≥1.21** | **CL ≥ 12** | **CL ≥ 24** | **CL ≥ 32** | **Cutpoint** | **Method** |
| p-tau217- \| Aβ-PET+ | 4 (1.2%) | 11 (3.5%) | 9 (2.9%) | 5 (1.6%) | 2 (0.7%) | 0.258 | Two-Point |
| p-tau217+ \| Aβ-PET- | 7 (2.1%) | 7 (2.2) | 8 (2.6%) | 10 (3.3%) | 11 (3.6%) | 0.472 | |
| p-tau217- \| Aβ-PET+ | 8 (2.4%) | 22 (7.0) | 18 (5.9%) | 9 (2.9%) | 5 (1.6%) | 0.338 | Binary (Youden) |
| p-tau217+ \| Aβ-PET- | 22 (6.7) | 22 (7.0) | 21 (6.8%) | 29 (9.4%) | 33 (10.7%) | 0.338 | |
| | *n* = 330 | *n* = 316 | *n* = 307 | | | | |

B

| | AB-PET POSITIVITY | | | | | ROC ANALYSES | |
|---|---|---|---|---|---|---|---|
| **Discordance** | **VISUAL READ** | **SUVR ≥ 1.21** | **CL ≥ 12** | **CL ≥ 24** | **CL ≥ 32** | **Cutpoint** | **Method** |
| p-tau217- \| Aβ-PET+ | 0 | 4 (1.88%) | 2 (.94%) | 0 | 0 | 0.258 | Two-Point |
| p-tau217+ \| Aβ-PET- | 6 (3.03%) | 6 (2.82%) | 5 (2.36%) | 7 (3.30%) | 8 (3.77%) | 0.472 | |
| p-tau217- \| Aβ-PET+ | 2 (1.01%) | 9 (4.23%) | 6 (2.83%) | 2 (.94%) | 2 (.94%) | 0.338 | Binary (Youden) |
| p-tau217+ \| Aβ-PET- | 14 (6.57%) | 14 (6.57%) | 12 (5.66%) | 18 (8.49%) | 21 (9.91%) | 0.338 | |
| | *n* = 218 | *n* = 214 | *n* = 213 | | | | |

**Table S5. Participant characteristics stratified by discordant status based on two-point and binary p-tau217 cutpoints using all available scans**

| | N | Overall N = 38[1] | p-tau217+/Aβ-PET- HIGH N = 10[1] | INT-HIGH N = 19[1] | p-tau217-/Aβ-PET+ INT-LOW N = 4[1] | LOW N = 5[1] |
|---|---|---|---|---|---|---|
| Dx | 38 | | | | | |
| CU | | 22 (58%) | 5 (50%) | 11 (58%) | 2 (50%) | 4 (80%) |
| MCI | | 14 (37%) | 3 (30%) | 8 (42%) | 2 (50%) | 1 (20%) |
| DEM | | 2 (5.3%) | 2 (20%) | 0 (0%) | 0 (0%) | 0 (0%) |
| Age (Baseline) | 38 | 71.7 (6.0) | 72.9 (5.6) | 70.4 (5.8) | 69.5 (6.9) | 76.2 (5.5) |
| Education | 38 | 16.50 (2.20) | 15.40 (3.03) | 17.37 (1.67) | 16.50 (1.00) | 15.40 (1.67) |
| Sex | 38 | 18 (47%) | 4 (40%) | 7 (37%) | 4 (100%) | 3 (60%) |
| Race | 38 | | | | | |
| Black | | 7 (18%) | 3 (30%) | 3 (16%) | 1 (25%) | 0 (0%) |
| White | | 31 (82%) | 7 (70%) | 16 (84%) | 3 (75%) | 5 (100%) |
| *APOE*-e4 Carrier | 37 | 13 (35%) | 4 (40%) | 6 (33%) | 2 (50%) | 1 (20%) |
| eGFR | 35 | 74 (14) | 74 (15) | 74 (13) | 82 (13) | 67 (17) |
| BMI | 38 | 28.8 (6.4) | 29.2 (5.8) | 28.5 (7.8) | 28.2 (3.1) | 29.4 (4.9) |
| CMI | 38 | -0.69 (0.65) | -0.69 (0.54) | -0.79 (0.73) | -0.38 (0.78) | -0.57 (0.40) |
| Insulin | 30 | 21 (17) | 12 (7) | 19 (13) | 40 (30) | 32 (21) |
| Glucose | 26 | 103 (21) | 93 (8) | 108 (26) | 106 (22) | 99 (NA) |
| Hemoglobin A1c | 25 | 5.68 (0.35) | 5.58 (0.39) | 5.63 (0.34) | 5.75 (0.49) | 5.92 (0.25) |
| HTN | 38 | 17 (45%) | 6 (60%) | 6 (32%) | 2 (50%) | 3 (60%) |
| HTNTX | 38 | 15 (39%) | 5 (50%) | 6 (32%) | 1 (25%) | 3 (60%) |
| Diabetes Group | 38 | | | | | |
| Diabetes | | 1 (2.6%) | 0 (0%) | 0 (0%) | 1 (25%) | 0 (0%) |
| Prediabetes | | 20 (53%) | 2 (20%) | 11 (58%) | 2 (50%) | 5 (100%) |
| p-tau217 | 38 | 0.45 (0.23) | 0.72 (0.29) | 0.39 (0.04) | 0.30 (0.03) | 0.24 (0.01) |
| Aβ-PET (SUVr) | 38 | 1.25 (0.19) | 1.15 (0.08) | 1.17 (0.11) | 1.59 (0.10) | 1.48 (0.15) |
| Aβ=PET (CL) | 38 | 12 (19) | 3 (9) | 4 (10) | 47 (12) | 35 (15) |
| Blood-PET Interval* | 38 | -529 (568) | -392 (548) | -384 (406) | -382 (251) | -1,474 (477) |

*Note*: Stratifying the cohort by self-reported discordance, *Abbreviations*: Aβ = Amyloid Beta; CMI = Cardiometabolic Index; HTN HX = History of Hypertension; eGFR = Estimated Glomerular Filtration Rate

**Table S6. Participant characteristics stratified by discordant status based on two-point and binary p-tau217 cutpoints for PET scans acquired withing 14 months from blood collection**

| | N | Overall N = 25[1] | p-tau217+/Aβ-PET- HIGH N = 8[1] | INT-HIGH N = 15[1] | p-tau217-/Aβ-PET+ INT-LOW N = 2[1] |
|---|---|---|---|---|---|
| Dx | 25 | | | | |
| CU | | 13 (52%) | 4 (50%) | 7 (47%) | 2 (100%) |
| DEM | | 1 (4.0%) | 1 (12%) | 0 (0%) | 0 (0%) |
| MCI | | 11 (44%) | 3 (38%) | 8 (53%) | 0 (0%) |
| Age (Baseline) | 25 | 71.2 (6.0) | 73.4 (6.1) | 70.5 (6.1) | 67.5 (3.5) |
| Education | 25 | 16.68 (2.34) | 15.75 (3.33) | 17.27 (1.71) | 16.00 (0.00) |
| Female | 25 | 13 (52%) | 4 (50%) | 7 (47%) | 2 (100%) |
| Race | 25 | | | | |
| Black | | 5 (20%) | 2 (25%) | 2 (13%) | 1 (50%) |
| White | | 20 (80%) | 6 (75%) | 13 (87%) | 1 (50%) |
| *APOE*-ε4 CARRIER | 24 | 9 (38%) | 3 (38%) | 5 (36%) | 1 (50%) |
| eGFR | 23 | 76 (11) | 80 (10) | 74 (12) | 77 (3) |
| BMI | 25 | 29 (7) | 30 (6) | 29 (9) | 30 (2) |
| CMI | 25 | -0.67 (0.71) | -0.71 (0.59) | -0.69 (0.75) | -0.37 (1.16) |
| Insulin | 20 | 19 (13) | 17 (13) | 19 (14) | 29 (NA) |
| Glucose | 20 | 104 (22) | 98 (17) | 105 (24) | 122 (NA) |
| Hemoglobin A1c | 14 | 5.60 (0.32) | 5.60 (0.42) | 5.60 (0.31) | NA (NA) |
| HTN | 25 | 10 (40%) | 4 (50%) | 6 (40%) | 0 (0%) |
| HTNTX | 25 | 8 (32%) | 3 (38%) | 5 (33%) | 0 (0%) |
| Diabetes Group | 25 | | | | |
| Diabetes | | 1 (4.0%) | 0 (0%) | 0 (0%) | 1 (50%) |
| Prediabetes | | 10 (40%) | 2 (25%) | 8 (53%) | 0 (0%) |
| p-tau217 | 25 | 0.47 (0.22) | 0.68 (0.30) | 0.38 (0.04) | 0.31 (0.01) |
| Aβ-PET (SUVr) | 25 | 1.22 (0.17) | 1.15 (0.10) | 1.20 (0.12) | 1.64 (0.04) |
| Aβ=PET (CL) | 25 | 10 (18) | 5 (12) | 7 (13) | 54 (1) |
| Blood-PET Interval* | 25 | -124 (101) | -84 (49) | -136 (113) | -195 (158) |

*Note*: Stratifying the cohort by self-reported discordance after restricting cases to those with less than 14 months between blood collection for p-tau217 and amyloid PET scan. *Abbreviations*: CMI = Cardiometabolic Index; HTN HX = History of Hypertension; eGFR = Estimated Glomerular Filtration Rate

**Table S7. Exploratory subgroup analysis of p-tau217 cutpoints before and after excluding large intervals between blood collection and scan acquisition**

| | | | | optimal | | | | | | max-sens | | | max-spec | | |
|---|---|---|---|---|---|---|---|---|---|---|---|---|---|---|---|
| | ***n*** | **auc** | **bootAUC** | **cp** | **bootCI** | **acc** | **sens** | **spec** | **prev** | **thresh** | **sens** | **npv** | **thresh** | **spec** | **ppv** |
| **Female** | 182 | .951 | .916-.978 | **.338** | .305-.412 | .901 | .889 | .908 | .346 | **.253** | .952 | .963 | **.400** | .950 | .895 |
| **Male** | 125 | .934 | .886-.975 | **.460** | .342-.496 | .904 | .857 | .928 | .336 | **.342** | .952 | .971 | **.546** | .952 | .882 |
| **White** | 260 | .941 | .910-.968 | **.341** | .332-.461 | .881 | .916 | .861 | .365 | **.253** | .947 | .952 | **.460** | .952 | .905 |
| **Black** | 42 | .929 | .831-.993 | **.305** | .305-.678 | .833 | 1.000 | .788 | .214 | **.286** | 1.00 | 1.00 | **.534** | .939 | .714 |
| **APOE-ε4-** | 208 | .930 | .889-.966 | **.322** | .322-.406 | .870 | .922 | .854 | .245 | **.241** | .941 | .967 | **.405** | .949 | .826 |
| ***APOE*-ε4+** | 91 | .940 | .878-.981 | **.491** | .370-.659 | .890 | .880 | .902 | .549 | **.368** | .940 | .917 | **.549** | .951 | .952 |
| ***eGFR < 60*** | 66 | .960 | .866-1 | **.509** | .508-.660 | .933 | .929 | .938 | .467 | **.489** | .929 | .938 | **.489** | ..938 | .929 |
| ***eGFR >= 60*** | 486 | .947 | .916-.970 | **.338** | .305-.412 | .890 | .907 | .882 | .326 | **.253** | .953 | .967 | **.431** | .950 | .880 |
| **AVERAGE** | | .942 | | **.388** | | .888 | .913 | .883 | .356 | **.311** | .952 | .959 | **.477** | .949 | .873 |

**Short Interval**

| | | | | optimal | | | | | | max-sens | | | max-spec | | |
|---|---|---|---|---|---|---|---|---|---|---|---|---|---|---|---|
| | ***n*** | **auc** | **bootAUC** | **cp** | **bootCI** | **acc** | **sens** | **spec** | **prev** | **thresh** | **sens** | **npv** | **thresh** | **spec** | **ppv** |
| **Female** | 125 | .969 | .940-.992 | **.338** | .305-.406 | .921 | .956 | .901 | .357 | **.338** | .956 | .973 | **.389** | .951 | .905 |
| **Male** | 87 | .969 | .927-.996 | **.460** | .359-.550 | .931 | .906 | .945 | .368 | **.371** | .938 | .960 | **.460** | .945 | .906 |
| **White** | 177 | .969 | .943-.989 | **.351** | .322-.461 | .910 | .971 | .872 | .384 | **.357** | .956 | .970 | **.460** | .954 | .921 |
| **Black** | 33 | .951 | .865-1 | **.305** | .305-.678 | .882 | 1.00 | .840 | .265 | **.282** | 1.00 | 1.00 | **.534** | .960 | .833 |
| ***APOE*-ε4-** | 67 | .950 | .887-.992 | **.550** | .370-.581 | .899 | .846 | .967 | .565 | **.371** | .949 | .923 | **.545** | .933 | .943 |
| ***APOE*-ε4+** | 132 | .970 | .942-.992 | **.338** | .322-.365 | .920 | .971 | .903 | .248 | **.340** | .941 | .979 | **.365** | .951 | .853 |
| ***eGFR < 60*** | 31 | 1 | N/A | **.508** | .508-.660 | .933 | .929 | 1 | .55 | **.551** | .910 | .900 | **.462** | 1 | 1 |
| ***eGFR >= 60*** | 101 | .901 | .824-.967 | **.443** | .234-.466 | .901 | .720 | .942 | .301 | **.235** | .960 | .533 | **.424** | .720 | .857 |
| **AVERAGE** | | **.960** | | **.412** | | **.912** | **.912** | **.921** | **.380** | **.356** | **.951** | **.905** | **.455** | **.927** | **.902** |

*Note:* Optimal ROC-based (e.g. binary; Youden) cutpoints are shown for each subgroup assessed in the current set of analyses before and after excluding large intervals between blood collection and scan acquisition (< 14 months). Variation across groups is captured within the bootstrapped confidence intervals and upper and lower detection thresholds. Stratified cutpoints are for descriptive purposes only; restricting analyses to individual subgroups artificially induces added sampling biases, and in most cases the number of observed cases is insufficient. Abbreviations: thresh= threshold (lower or upper boundary); sens = sensitivity; spec = specificity; ppv = positive predictive value; npv = negative predictive values

**Table S8. Primary variables stratified by Self-Reported Sex**

| | N | **Overall** (*n*=598)[1] | **Female** (n=388)[1] | **Male** (n=210)[1] | **p**[2] | **ES** |
|---|---|---|---|---|---|---|
| **Age** | 598 | 70 (8) | 69 (8) | 71 (8) | **.041** | **.084** |
| **Education (years)** | 598 | 15.81 (2.55) | 15.59 (2.48) | 16.21 (2.65) | **.004** | **.116** |
| **Race: Black** | 598 | 124 (21%) | 99 (26%) | 25 (12%) | **<.001** | **.180** |
| *APOE*-ε4 | 579 | 187 (32%) | 120 (32%) | 67 (34%) | .600 | .017 |
| eGFR | 552 | 78 (15) | 78 (16) | 77 (13) | .110 | .067 |
| BMI | 598 | 27.9 (5.6) | 27.6 (5.8) | 28.3 (5.3) | .070 | .074 |
| **CMI** | 587 | -0.82 (0.50) | -0.79 (0.49) | -0.87 (0.51) | **.029** | **.090** |
| Insulin | 484 | 25 (31) | 24 (32) | 25 (30) | .500 | .034 |
| **Glucose** | 413 | 104 (24) | 102 (24) | 108 (24) | **<.001** | **.166** |
| Hemoglobin A1c | 373 | 5.71 (0.51) | 5.69 (0.49) | 5.75 (0.53) | .200 | .074 |
| **HTN HX** | 598 | 260 (43%) | 152 (39%) | 108 (51%) | **.004** | **.114** |
| Pre/Diabetes | 590 | 358 | 233 | 125 | .800 | .026 |
| p-tau181 | | | | | | |
| **p-tau217** | 598 | 0.44 (0.36) | 0.42 (0.36) | 0.47 (0.36) | **.007** | **.109** |
| SUVr | 316 | 1.39 (0.44) | 1.39 (0.45) | 1.38 (0.42) | .600 | .027 |
| Centiloids | 307 | 27 (44) | 28 (45) | 26 (42) | >0.9 | .005 |
| Blood-PET Interval | 330 | -383 (624) | -382 (644) | -384 (594) | .700 | .019 |

*Note*: Stratifying the cohort by self-reported sex, Female participants were, on average, slightly younger and less educated, had a higher proportion of Black participants, worse kidney functioning, lower blood glucose levels, history of hypertension, and p-tau217 levels.
*Abbreviations*: CMI = Cardiometabolic Index; HTN HX = History of Hypertension; eGFR = Estimated Glomerular Filtration Rate

**Table S9. Primary variables stratified by Self-Reported Race**

| | N | Overall (n=591)[1] | Black (n=124)[1] | White (n=467)[1] | p[2] | EF |
|---|---|---|---|---|---|---|
| **Age** | **591** | **70 (8)** | **68 (8)** | **70 (8)** | **.003** | **.123** |
| **Education** | **591** | **15.81 (2.54)** | **15.25 (2.49)** | **15.96 (2.54)** | **.008** | **.109** |
| **Sex: Female** | **591** | **384 (65%)** | **99 (80%)** | **285 (61%)** | **<.001** | **.156** |
| ***APOE*-ε4** | **572** | **185 (32%)** | **49 (41%)** | **136 (30%)** | **.025** | **.089** |
| **eGFR** | **545** | **78 (15)** | **73 (16)** | **79 (14)** | **<.001** | **.176** |
| **BMI** | **591** | **27.9 (5.6)** | **30.7 (6.3)** | **27.1 (5.2)** | **<.001** | **.246** |
| **CMI** | **580** | **-0.82 (0.50)** | **-0.61 (0.51)** | **-0.87 (0.48)** | **<.001** | **.216** |
| **Insulin** | **480** | **24 (31)** | **30 (43)** | **23 (27)** | **.039** | **.094** |
| Glucose | 410 | 104 (24) | 105 (27) | 104 (23) | >.900 | .001 |
| **Hemoglobin A1c** | **367** | **5.71 (0.51)** | **5.95 (0.59)** | **5.65 (0.46)** | **<.001** | **.281** |
| Pre/Diabetes | 583 | 353 (61%) | 83 (67%) | 270 (59%) | .100 | .064 |
| **HTN HX** | **591** | **258 (44%)** | **80 (65%)** | **178 (38%)** | **<.001** | **.213** |
| p-tau181 | 530 | 3.47 (2.00) | 3.35 (2.19) | 3.50 (1.96) | .130 | .065 |
| **p-tau217** | **591** | **0.44 (0.36)** | **0.33 (0.26)** | **0.47 (0.38)** | **<.001** | **.196** |
| **SUVr** | **311** | **1.39 (0.44)** | **1.22 (0.31)** | **1.42 (0.46)** | **.001** | **.182** |
| Centiloids | 302 | 27 (44) | 13 (31) | 30 (45) | .058 | .109 |
| Blood-PET Interval | 325 | -379 (624) | -231 (551) | -402 (633) | .200 | .070 |

*Note*: Stratifying the cohort by self-reported race, Black participants were, on average, slightly younger and less educated, had a higher proportion of female participants, worse kidney functioning, poorer cardiometabolic health (higher BMI, CMI, Insulin, and A1c), history of hypertension, elevated p-tau217 and amyloid PET (SUVr) levels. *Abbreviations*: CMI = Cardiometabolic Index; HTN HX = History of Hypertension; eGFR = Estimated Glomerular Filtration Rate

**Table S10. Primary variables stratified by *APOE*-ε4 status**

| | n | Overall (n= 572)[1] | CARRIER (n=185)[1] | NONCARRIER (n=387)[1] | p[2] | EF |
|---|---|---|---|---|---|---|
| **Age** | **572** | **70 (8)** | **68 (7)** | **71 (8)** | **0.003** | **0.123** |
| Education | 572 | 15.83 (2.55) | 15.89 (2.54) | 15.80 (2.56) | 0.7 | 0.014 |
| Sex: Female | 572 | 376 (66%) | 119 (64%) | 257 (66%) | 0.6 | |
| **Race: Black** | **572** | **120 (21%)** | **49 (26%)** | **71 (18%)** | **0.025** | |
| eGFR | 529 | 78 (15) | 78 (15) | 78 (14) | 0.9 | 0.008 |
| BMI | 572 | 27.9 (5.6) | 27.4 (5.5) | 28.1 (5.7) | 0.11 | 0.067 |
| CMI | 565 | -0.82 (0.50) | -0.84 (0.47) | -0.81 (0.51) | 0.7 | 0.016 |
| Insulin | 466 | 25 (32) | 26 (38) | 24 (28) | 0.6 | 0.025 |
| Glucose | 396 | 104 (24) | 103 (23) | 105 (25) | 0.3 | 0.049 |
| Hemoglobin A1c | 361 | 5.72 (0.51) | 5.66 (0.38) | 5.74 (0.56) | 0.4 | 0.044 |
| Pre/Diabetes | 567 | 347 (61%) | 111 (60%) | 236 (62%) | 0.8 | |
| HTN HX | 572 | 248 (43%) | 76 (41%) | 172 (44%) | 0.4 | |
| **p-tau181** | **515** | **3.44 (1.98)** | **3.68 (1.72)** | **3.33 (2.08)** | **.001** | **0.142** |
| **p-tau217** | **572** | **0.43 (0.36)** | **0.54 (0.40)** | **0.38 (0.32)** | **<.001** | **0.226** |
| **SUVr** | **303** | **1.38 (0.44)** | **1.59 (0.50)** | **1.29 (0.38)** | **<.001** | **0.278** |
| **Centiloids** | **294** | **27 (44)** | **50 (49)** | **17 (37)** | **<.001** | **0.316** |
| Blood-PET Interval | 317 | -386 (630) | -338 (594) | -408 (646) | 0.5 | 0.037 |

*Note*: Stratifying the cohort by *APOE*-ε4 status, *APOE*-ε4 carriers were, on average, slightly younger, had a higher proportion of Black participants elevated p-tau181 and p-tau217 levels. *Abbreviations*: CMI = Cardiometabolic Index; HTN HX = History of Hypertension; eGFR = Estimated Glomerular Filtration Rate

#### Table S11. Primary variables stratified by eGFR

| | n | **Overall**, N = 552[1] | **Stage 3**, N = 66[1] | **Stage 1-2**, N = 486[1] | **p-value**[2] | EF |
|---|---|---|---|---|---|---|
| **Age** | **552** | **70 (8)** | **75 (8)** | **69 (8)** | **<0.001** | **0.215** |
| **Education** | **552** | **15.83 (2.55)** | **15.05 (2.71)** | **15.93 (2.51)** | **0.01** | **0.109** |
| Sex: Female | 552 | 363 (66%) | 45 (68%) | 318 (65%) | 0.7 | 0.013 |
| Race: Black | 552 | 114 (21%) | 22 (33%) | 92 (19%) | 0.062 | 0.121 |
| *APOE*-ε4 | 536 | 176 (33%) | 19 (31%) | 157 (33%) | 0.8 | 0.007 |
| **BMI** | **552** | **27.9 (5.6)** | **29.3 (5.7)** | **27.7 (5.6)** | **0.008** | **0.113** |
| CMI | 543 | -0.82 (0.50) | -0.78 (0.43) | -0.82 (0.51) | 0.3 | 0.047 |
| **Insulin** | **484** | **25 (31)** | **33 (49)** | **23 (28)** | **0.002** | **0.143** |
| Glucose | 413 | 104 (24) | 107 (24) | 104 (24) | 0.2 | 0.062 |
| hemoglobin_a1c | 373 | 5.71 (0.51) | 5.77 (0.42) | 5.71 (0.52) | 0.2 | 0.06 |
| Pre/Diabetes | 544 | 332 (61%) | 43 (67%) | 289 (60%) | 0.3 | 0.04 |
| **HTN HX** | **552** | **241 (44%)** | **46 (70%)** | **195 (40%)** | **<0.001** | **0.188** |
| **p-tau181** | **495** | **3.41 (1.98)** | **4.29 (2.26)** | **3.28 (1.91)** | **<0.001** | **0.195** |
| **p-tau217** | **552** | **0.43 (0.35)** | **0.63 (0.51)** | **0.40 (0.31)** | **<0.001** | **0.17** |
| suvr1_read | 303 | 1.38 (0.44) | 1.51 (0.51) | 1.37 (0.43) | 0.1 | 0.094 |
| Centiloids | 294 | 27 (44) | 39 (49) | 25 (43) | 0.11 | 0.092 |
| Blood-PET Interval | 317 | -394 (633) | -423 (584) | -391 (639) | 0.9 | 0.01 |

[1] n (%); Mean (SD)

[2] Fisher's exact test; Wilcoxon rank sum test; Pearson's Chi-squared test

*Note*: Stratifying the cohort by eGFR (eGFR < 60 and eGFR > 60), participants with an eGFR < 60 are, on average, slightly older and lower educated, and with a current or recent history of hypertension, and have higher p-tau181 and p-tau217 levels. As 60 represents a somewhat arbitrary cutpoint, we also looked at typical stages (see Table S12). *Abbreviations*: CMI = Cardiometabolic Index; HTN HX = History of Hypertension; eGFR = Estimated Glomerular Filtration Rate; *APOE*-ε4

**Table S12. Primary variables stratified by eGFR Stages**

| | **N** | **Overall, N = 552[1]** | **Stage1, N = 135[1]** | **Stage2, N = 351[1]** | **Stage3, N = 66[1]** | **p[2]** |
|---|---|---|---|---|---|---|
| **Age** | **552** | **70 (8)** | **66 (6)** | **70 (8)** | **75 (8)** | **<0.001** |
| **Education** | **552** | **15.83 (2.55)** | **15.69 (2.48)** | **16.03 (2.52)** | **15.05 (2.71)** | **0.015** |
| Sex: Female | 552 | 363 (66%) | 96 (71%) | 222 (63%) | 45 (68%) | 0.2 |
| **Race: Black** | **552** | **114 (21%)** | **17 (13%)** | **75 (21%)** | **22 (33%)** | **0.02** |
| *APOE*-ε4 | 536 | 176 (33%) | 48 (36%) | 109 (32%) | 19 (31%) | 0.6 |
| **BMI** | **552** | **27.9 (5.6)** | **27.2 (5.4)** | **27.9 (5.6)** | **29.3 (5.7)** | **0.018** |
| CMI | 543 | -0.82 (0.50) | -0.81 (0.51) | -0.82 (0.51) | -0.78 (0.43) | 0.5 |
| **Insulin** | **484** | **25 (31)** | **21 (22)** | **24 (30)** | **33 (49)** | **0.007** |
| Glucose | 413 | 104 (24) | 104 (22) | 104 (25) | 107 (24) | 0.4 |
| Hemoglobin A1c | 373 | 5.71 (0.51) | 5.73 (0.54) | 5.70 (0.51) | 5.77 (0.42) | 0.5 |
| Pre/Diabetes | 544 | 332 (61%) | 84 (63%) | 205 (59%) | 43 (67%) | 0.4 |
| **HTN HX** | **552** | **241 (44%)** | **56 (41%)** | **139 (40%)** | **46 (70%)** | **<0.001** |
| **p-tau181** | **495** | **3.41 (1.98)** | **2.81 (1.33)** | **3.47 (2.07)** | **4.29 (2.26)** | **<0.001** |
| **p-tau217** | **552** | **0.43 (0.35)** | **0.34 (0.27)** | **0.42 (0.33)** | **0.63 (0.51)** | **<0.001** |
| SUVr | 303 | 1.38 (0.44) | 1.32 (0.39) | 1.38 (0.44) | 1.51 (0.51) | 0.2 |
| Centiloids | 294 | 27 (44) | 21 (39) | 27 (44) | 39 (49) | 0.3 |
| Blood-PET Interval | 317 | -394 (633) | -473 (602) | -360 (651) | -423 (584) | 0.3 |

[1] n (%); Mean (SD); [2] Pearson's Chi-squared test; Fisher's exact test;

*Note*: Stratifying the cohort by eGFR stages present within the Wake Forest ADRC Clinical Cohort, those in more developed stages (2-3) are, on average, slightly older and lower educated, comprised of a higher proportion of Black participants and with a current or recent history of hypertension, and have higher p-tau181 and p-tau217 levels. Stage of kidney disease estimated by eGFR (Stage 1 > 60; Stage 2 = 60-90; Stage 3 < 60). *Abbreviations*: CMI = Cardiometabolic Index; HTN HX = History of Hypertension; eGFR = Estimated Glomerular Filtration Rate

**Table S13. Estimated costs & savings for prioritizing scans**

| Single Follow-Up (PET Scan Costs Only) | # Scans | Amyloid $2,050 | Tau $1,600 | TOTAL $3,650 | % Savings Total | % Savings Year |
|---|---|---|---|---|---|---|
| **Total** | 598 | 1226k | 957k | **$2,182,700** | -- | -- |
| **Per Year** | 140 | 287k | 224k | **511k** | **76.6** | **--** |
| **Zone 3** | 109 | 223k | 0k | **223k** | **89.8** | **56.3** |
| **Zone 2** | 77 | 0k | 123k | **123k** | **94.4** | **75.9** |
| **Zone 1** | 171 | 0k | 274k | **274k** | **87.5** | **46.5** |
| **Revised Totals** | **357** | **223450** | **396800** | **620k** | | |

*Note:* Development of a research-oriented priority scan system would require fewer confirmatory Aβ-PET scans (Mattsson-Carlgren et al., 2023) and to prioritize initial and follow-up longitudinal tau-PET. Current estimates suggest ~5 years on average for tau-accumulation post amyloid onset (Josephs et al., 2022), though the rate of accumulation in diverse and heterogeneous cohorts is uncertain and is likely impacted by demographics, comorbidities, and other health-related conditions (Landau et al., 2024; Ossenkoppele et al., 2020; Smith et al., 2020). *Abbreviations*: Zone 1 (Absolute Positive) = p-tau217 ≥ .472 pg/mL; Zone 2 (Intermediate-Positive) = p-tau217 ≥ .338 pg/mL and p-tau217 < .472 pg/mL; Zone 3 (Intermediate-Negative) = p-tau217 ≥ .253 pg/mL and p-tau217 < ..338 pg/mL

#### 3 SUPPLEMENTAL FIGURES

**Figure S1. Gaussian-mixture model-based plasma p-tau217 cutpoints**

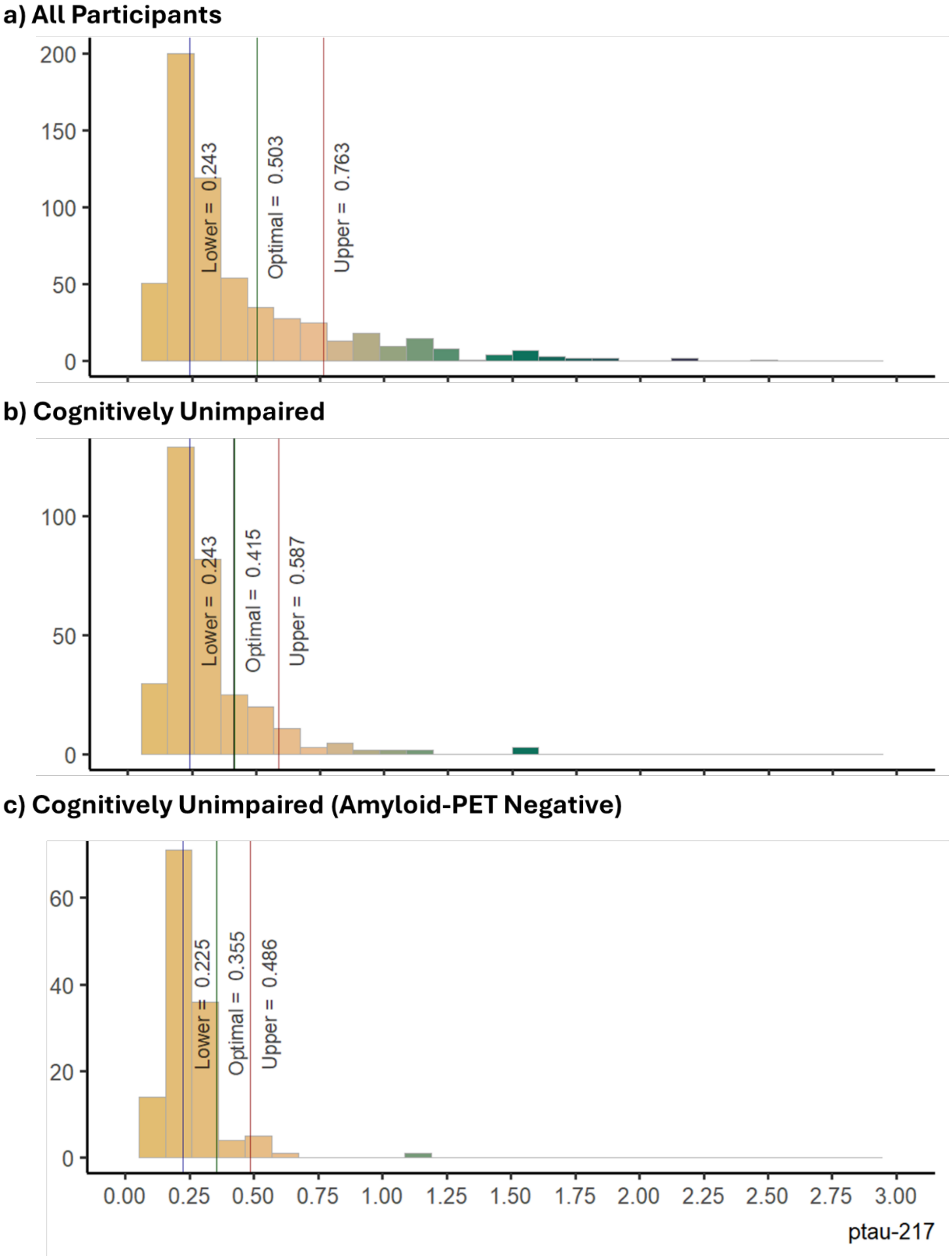


*Note:* Cross-sectional plasma p-tau217 cutpoints derived using gaussian-mixture modeling (GMM; outcome agnostic [e.g., irrespective of amyloid-PET status]) are shown in (panel a) all participants, (panel b) cognitively unimpaired (CU) participants, and (panel c) CU amyloid-PET negative participants. GMM-based cutpoints.

**Figure S2. Time interval between blood collection and amyloid PET scan at baseline**

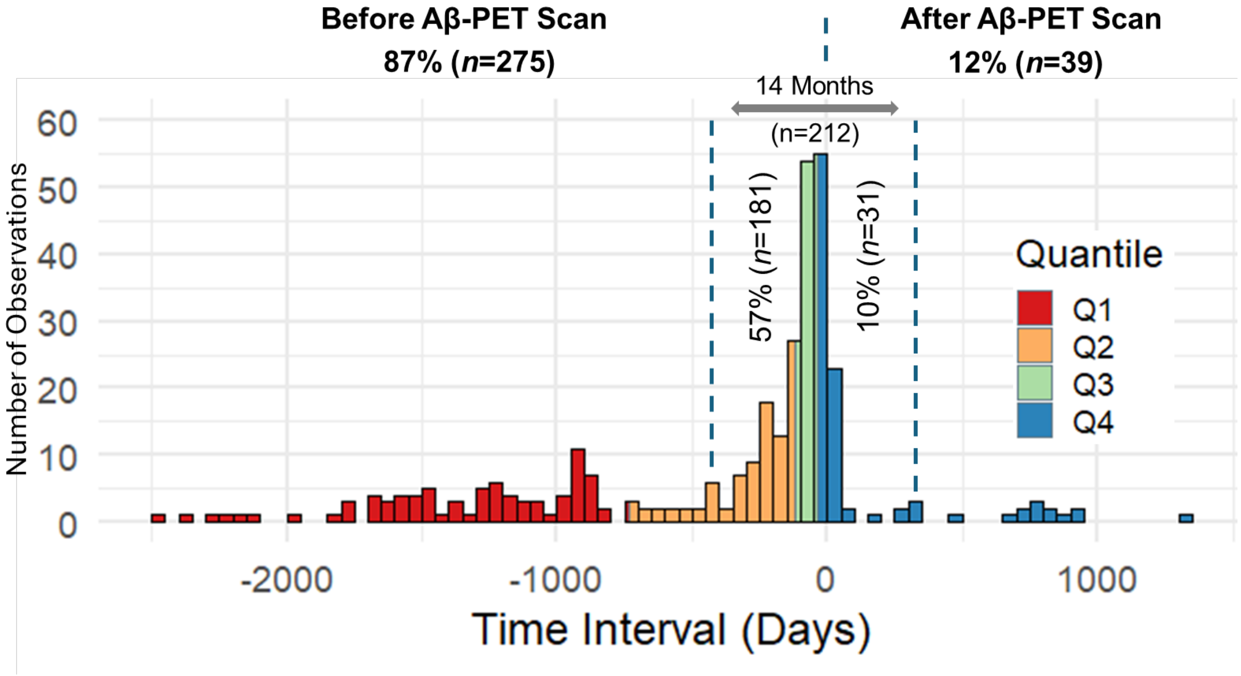

**Figure S3. P-tau217 cutpoints derived from the entire cohort superimposed on datapoints stratified by clinical diagnosis**

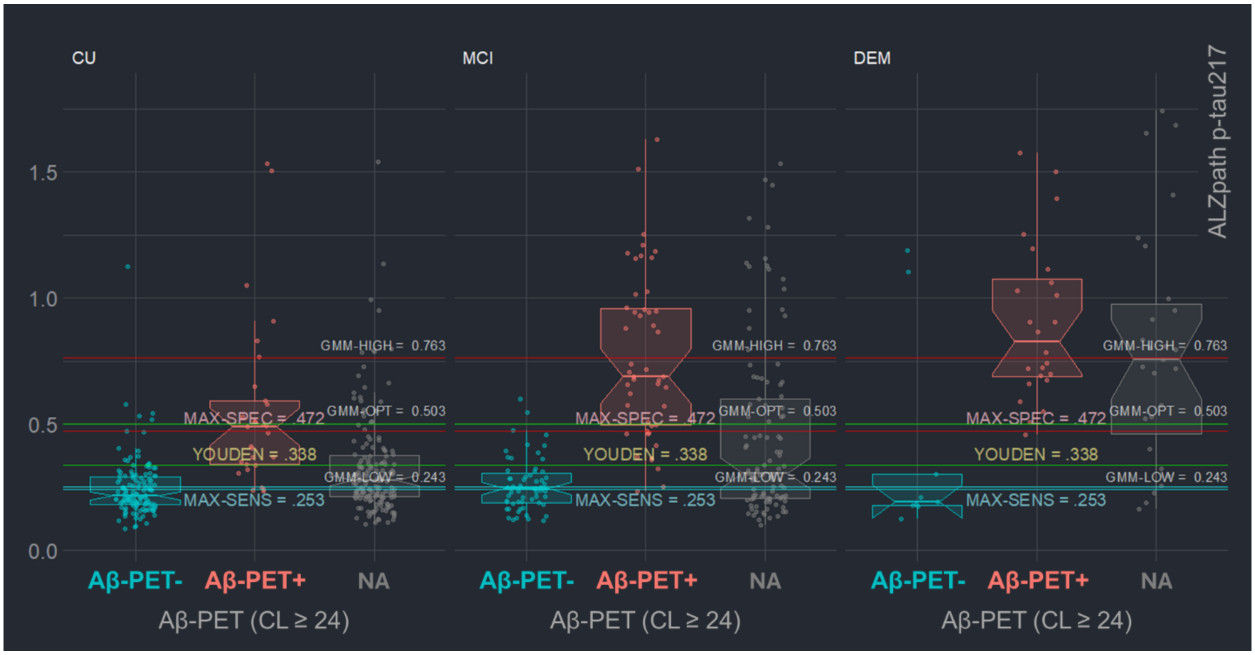

**Figure S4. P-tau217 cutpoints derived from the entire cohort superimposed on datapoints stratified by sex**

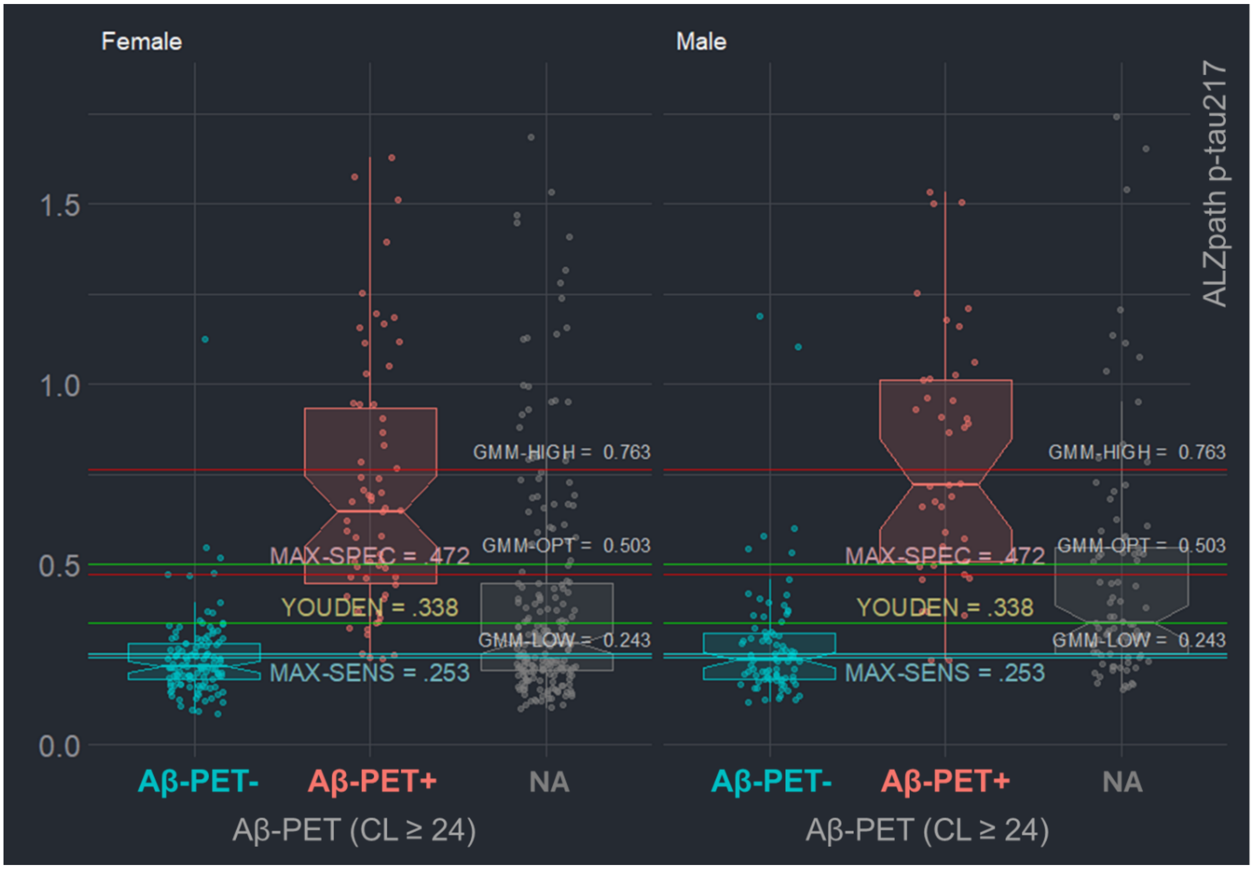

**Figure S5. P-tau217 cutpoints derived from the entire cohort superimposed on datapoints stratified by race**

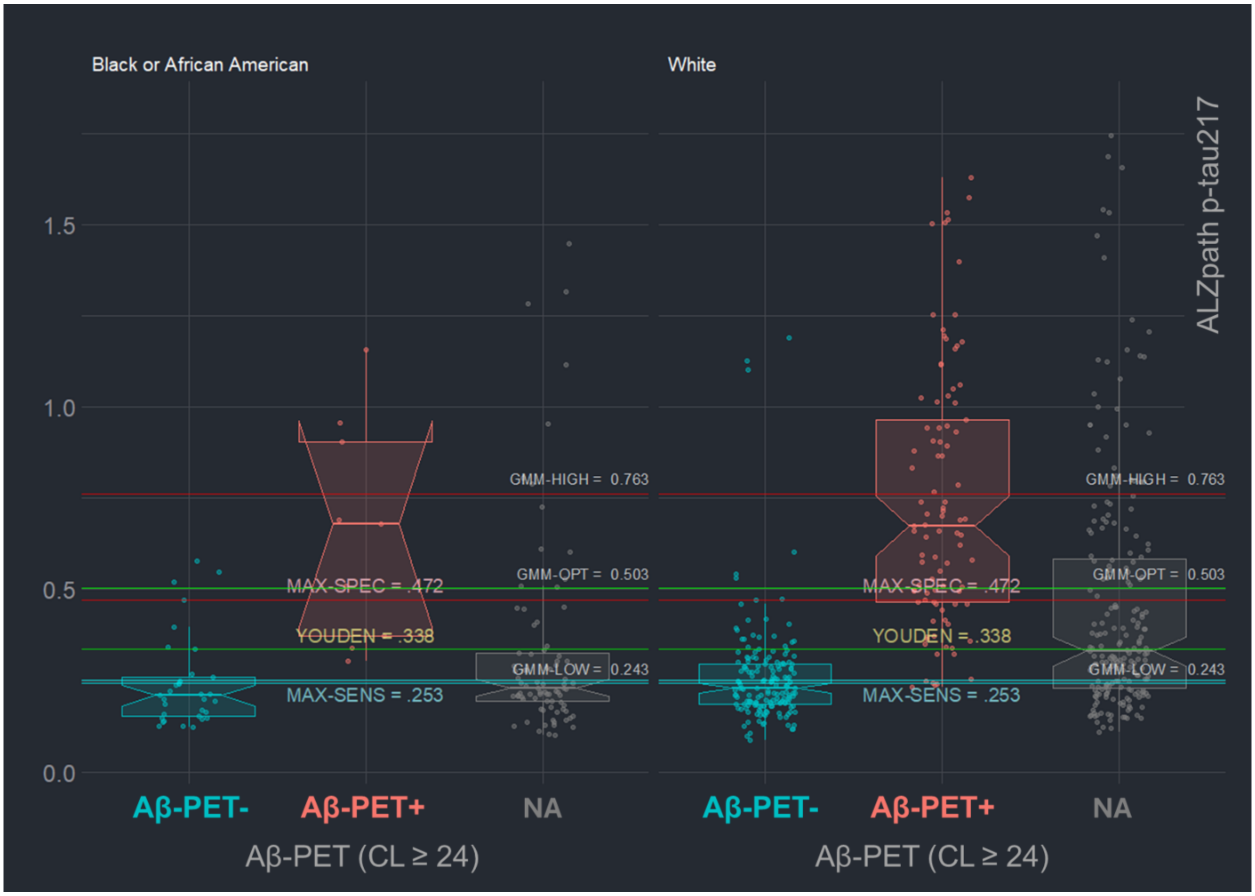

**Figure S6. P-tau217 cutpoints derived from the entire cohort superimposed on datapoints stratified by APOE-ε4 carriership**

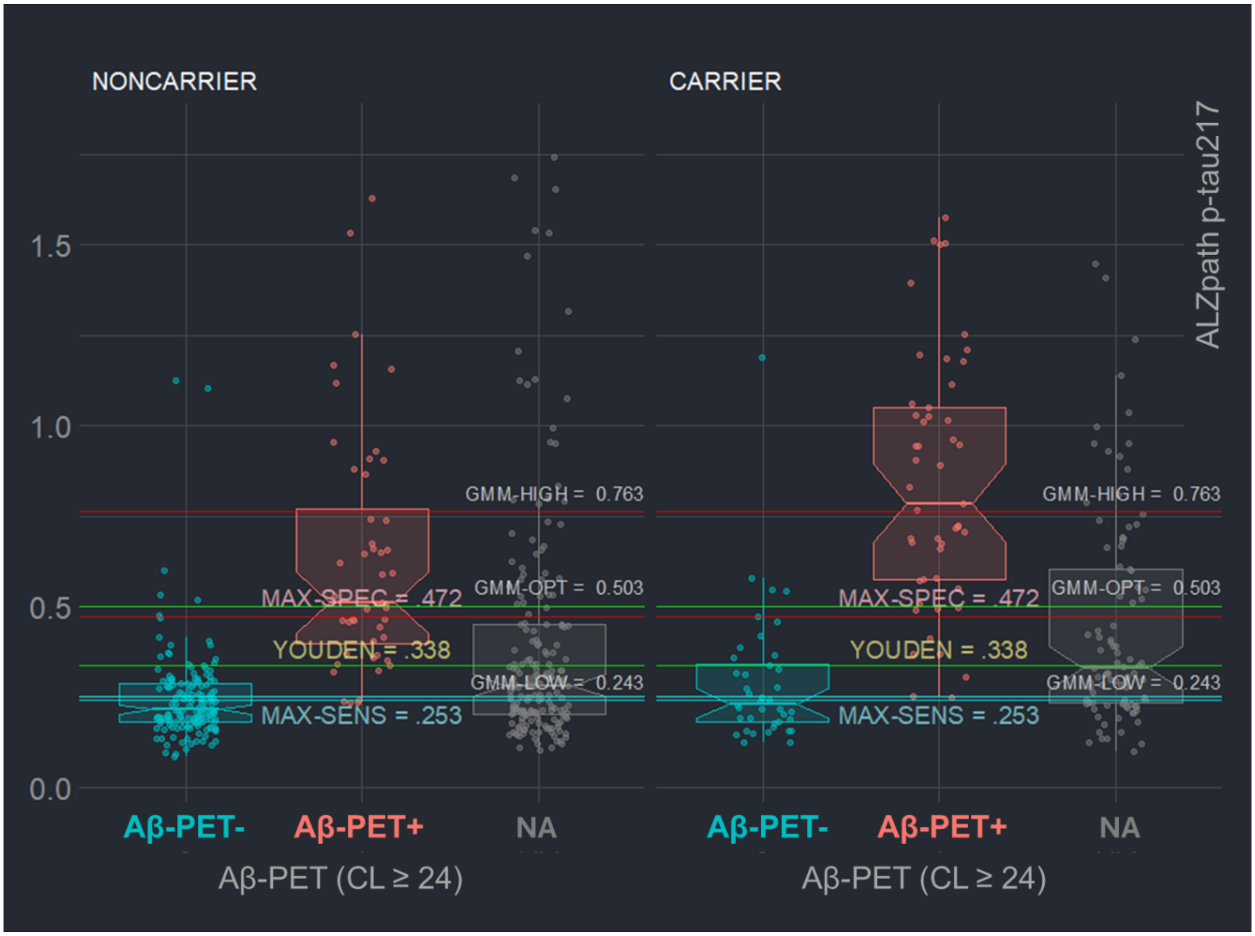

**Figure S7. P-tau217 cutpoints derived from the entire cohort superimposed on datapoints stratified by kidney functioning**

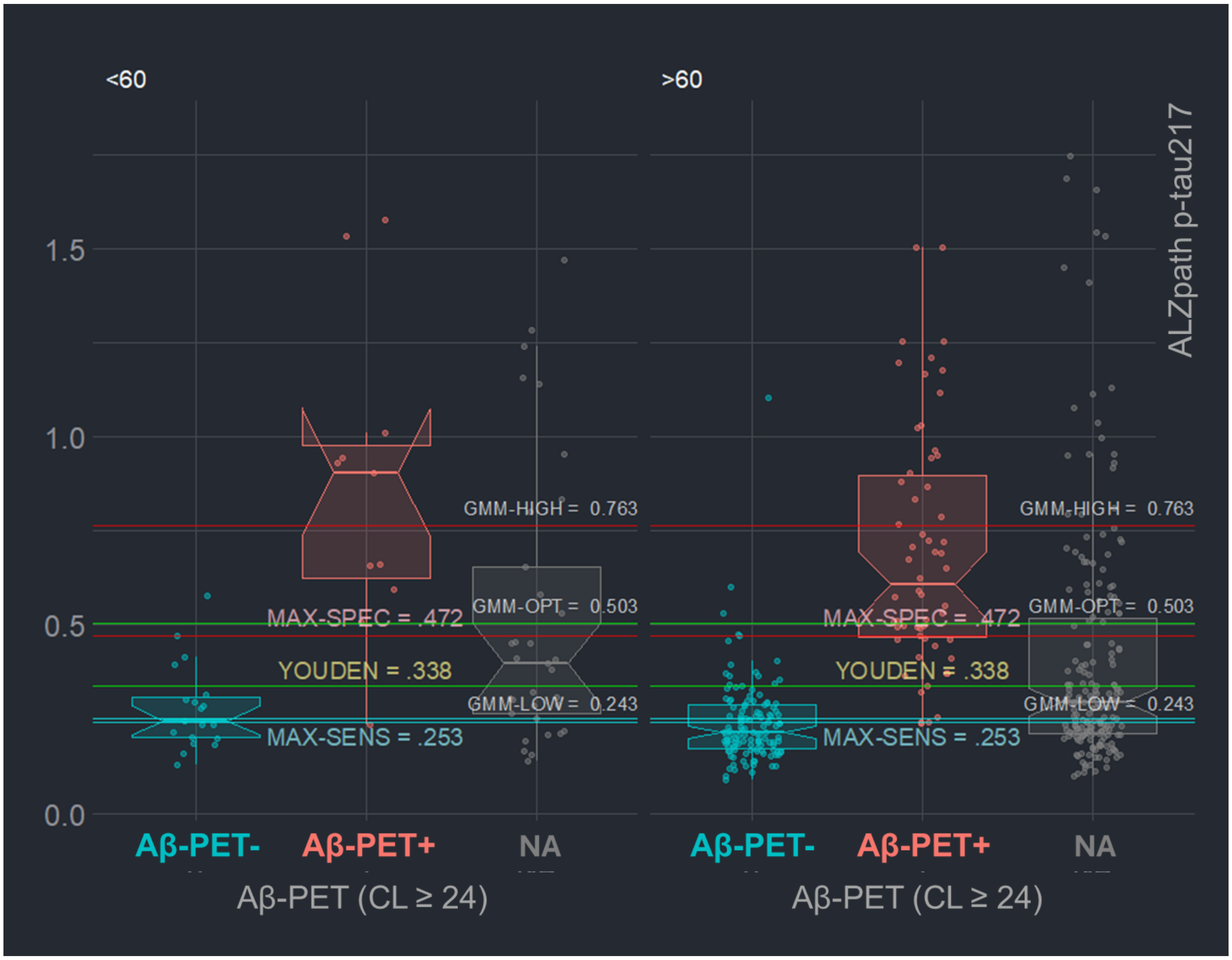

**SUPPLEMENTAL REFERENCES**


Hughes, T. M., Lockhart, S. N., Suerken, C. K., Jung, Y., Whitlow, C. T., Bateman, J. R., Williams, B. J., Espeland, M. A., Sachs, B. C., Williamson, J., Cleveland, M., Yang, M., Rogers, S., Hayden, K. M., Baker, L. D., & Craft, S. (2022). Hypertensive Aspects of Cardiometabolic Disorders Are Associated with Lower Brain Microstructure, Perfusion, and Cognition. *Journal of Alzheimer's Disease : JAD*, *90*(4), 1589. https://doi.org/10.3233/JAD-220646

Josephs, K. A., Weigand, S. D., & Whitwell, J. L. (2022). Characterizing Amyloid-Positive Individuals With Normal Tau PET Levels After 5 Years: An ADNI Study. *Neurology*, *98*(22), E2282–E2292. https://doi.org/10.1212/WNL.0000000000200287

Landau, S. M., Lee, J. Q., Murphy, A., Ward, T. J., Harrison, T. M., Baker, S. L., DeCarli, C., Harvey, D., Tosun, D., Weiner, M. W., Koeppe, R. A., & Jagust, W. J. (2024). Individuals with Alzheimer's disease and low tau burden: Characteristics and implications. *Alzheimer's & Dementia*, *20*(3), 2113–2127. https://doi.org/10.1002/ALZ.13609

Mattsson-Carlgren, N., Collij, L. E., Stomrud, E., Pichet Binette, A., Ossenkoppele, R., Smith, R., Karlsson, L., Lantero-Rodriguez, J., Snellman, A., Strandberg, O., Palmqvist, S., Ashton, N. J., Blennow, K., Janelidze, S., Hansson, O., & Authors, C. (2023). Plasma Biomarker Strategy for Selecting Patients With Alzheimer Disease for Antiamyloid Immunotherapies. *JAMA Neurology*. https://doi.org/10.1001/JAMANEUROL.2023.4596

Ossenkoppele, R., Leuzy, A., Cho, H., Sudre, C. H., Strandberg, O., Smith, R., Palmqvist, S., Mattsson-Carlgren, N., Olsson, T., Jögi, J., Stormrud, E., Ryu, Y. H., Choi, J. Y., Boxer, A. L., Gorno-Tempini, M. L., Miller, B. L., Soleimani-Meigooni, D., Iaccarino, L., La Joie, R., … Hansson, O. (2020). The impact of demographic, clinical, genetic, and imaging variables on tau PET status. *European Journal of Nuclear Medicine and Molecular Imaging*, *48*(7), 2245. https://doi.org/10.1007/S00259-020-05099-W

Rudolph, M. D., Sutphen, C. L., Register, T. C., Whitlow, C. T., Solingapuram Sai, K. K., Hughes, T. M., Bateman, J. R., Dage, J. L., Russ, K. A., Mielke, M. M., Craft, S., & Lockhart, S. N. (2024). Associations among plasma, MRI, and amyloid PET biomarkers of Alzheimer's disease and related dementias and the impact of health-related comorbidities in a community-dwelling cohort. *Alzheimer's & Dementia*, *20*(6), 4159. https://doi.org/10.1002/ALZ.13835

Smith, R., Strandberg, O., Mattsson-Carlgren, N., Leuzy, A., Palmqvist, S., Pontecorvo, M. J., Devous, M. D., Ossenkoppele, R., & Hansson, O. (2020). The accumulation rate of tau aggregates is higher in females and younger amyloid-positive subjects. *Brain*, *143*(12), 3805–3815. https://doi.org/10.1093/BRAIN/AWAA327
